# Physiology-Informed Conditional Variational Autoencoder for Generating Pediatric Virtual Patients

**DOI:** 10.64898/2026.01.21.26344442

**Authors:** Kei Irie, Tomoyuki Mizuno

**Affiliations:** Division of Translational and Clinical Pharmacology, Cincinnati Children’s Hospital Medical Center, Cincinnati, Ohio, USA; Department of Pediatrics, University of Cincinnati College of Medicine, Cincinnati, Ohio, USA

## Abstract

Reliable pediatric virtual patients are essential for model-informed simulations, including physiologically based pharmacokinetic (PBPK) modeling, to support dose selections in children and to evaluate drug exposure across developmental stages. Despite the availability of extensive pediatric physiological data and age- or size-based models, there remains a lack of well-established, flexible, and scalable approaches for integrating these data into realistic pediatric virtual patients that preserve multivariate physiological correlations and whole-body coherence across diverse clinical conditions and population needs.

In this proof-of-concept study, we developed a physiology-informed conditional variational autoencoder (cVAE) to address this challenge. The model was trained using real-world pediatric data augmented with mechanistically derived physiological information and conditioned on age and sex. It generated realistic physiological parameters, including body size, estimated glomerular filtration rate, organ weights, and blood flows, while biological plausibility was maintained through embedded physiological constraints.

The trained model demonstrated high reconstruction accuracy, with a mean absolute error of 0.0043 and an R² of 0.998, and the generated distributions closely matched those of the training data. All synthesized physiological profiles satisfied predefined physiological constraints, with total organ mass remaining below body weight and the sum of organ blood flows not exceeding cardiac output. Latent-space analyses further revealed smooth developmental patterns, enabling targeted physiological profile generation. The applicability of the generated physiological data was further demonstrated through PBPK simulations conducted across the pediatric age range using vancomycin as a testbed.

Overall, this physiology-informed generative framework supports coherent pediatric virtual patient generation for PBPK modeling and model-informed dosing applications development.

**Study Highlights:** *What is the current knowledge on the topic?:* Model-informed simulation, including physiologically based pharmacokinetic (PBPK) modeling, provides useful estimation of pediatric drug disposition across developmental stages and supports pediatric dose selection. However, constructing physiologically coherent pediatric virtual populations remains challenging. Although real-world pediatric measurements and physiologically derived, function-based information are available, these data are typically obtained from heterogeneous sources. Integrating them to generate multivariate physiological profiles at the individual level, while maintaining internal coherence across interconnected organ systems, remains an open challenge in pediatric pharmacometric modeling.

*What question did this study address?:* Can a conditional generative modeling and latent representation learning framework that integrates real-world pediatric data with mechanistically derived physiological constraints generate biologically coherent, multivariate pediatric physiological profiles that are suitable for downstream PBPK modeling across the full pediatric age range?

*What does this study add to our knowledge?:* This study introduces a physiology-informed conditional variational autoencoder that learns a smooth, interpretable latent space of pediatric physiology conditioned on age and sex. By embedding physiological constraints directly into the training objective, the model generates virtual pediatric patients with internally consistent body size, renal function, organ weights, and blood flows. The utility of these generated profiles was demonstrated through latent-space inversion and vancomycin PBPK simulations that reproduced reported age-dependent exposure trends and variability.

*How might this change clinical pharmacology or translational science?:* This framework provides a scalable approach for generating physiologically coherent pediatric virtual populations, laying a foundation for robust and flexible mechanistic PK simulations, virtual clinical trials, and digital twin applications. It offers a practical bridge between real-world pediatric data and mechanistic modeling, supporting model-informed dosing and translational decision-making in pediatric patient care and drug development.

## INTRODUCTION

Providing safe, effective, and evidence-based medication to children remains one of the most pressing challenges in clinical pharmacology and global health. Despite advances in precision medicine, pediatric patients are often underrepresented in clinical trials, and many medications are prescribed off-label, often without rigorous efficacy and safety data to guide optimal dosing (1). This lack of tailored evidence is particularly concerning, given that children undergo rapid and nonlinear developmental changes that can profoundly affect drug response, risk of toxicity, and variable treatment outcomes (2).

Physiologically based pharmacokinetic (PBPK) modeling has emerged as a key approach for addressing these gaps. By integrating organ volumes, blood flows, developmental physiology, and drug-specific properties into a mechanistic framework, PBPK models can predict how pharmacokinetics change with age and body size, simulate exposure under different dosing regimens, and extrapolate from adult or older pediatric data to younger age groups (3–6). In this way, PBPK modeling supports age-appropriate dose prediction and evaluation of alternative dosing strategies, particularly in settings where prospective pediatric studies are ethically or logistically constrained. Accurate and developmentally appropriate physiological information is therefore essential for generating reliable pediatric PBPK predictions and for using these models as quantitative evidence to inform dosing decisions (7, 8).

Over the past decades, large pediatric datasets covering wide age ranges have become available for variables such as body weight, height, and basic laboratory measurements, enabling characterization of population-level growth and developmental trends (9). In contrast, physiological parameters that are difficult to measure directly, including organ weights and tissue-specific blood flows, have generally been derived from heterogeneous sources using a variety of empirical observations, mechanistic considerations, and scaling assumptions. To represent these organ-level parameters across the pediatric age spectrum, age- or size-dependent functions are often used to describe developmental changes in physiology (10, 11).

While these approaches provide useful approximations of pediatric physiology, integrating real-world pediatric measurements with physiologically derived, function-based information to construct multivariate physiological profiles remains challenging. In particular, ensuring internal coherence across multiple organ systems at the individual level can be difficult when combining data-driven observations with mechanistic scaling relationships originating from different sources. Consequently, harmonizing heterogeneous physiological information into a coherent representation of an individual child remains an open challenge. Flexible and scalable approaches for generating physiologically coherent representations are increasingly important for advancing model-informed simulations, including extensions of physiologically based pharmacokinetic frameworks toward digital-twin concepts and large-scale in silico pediatric studies, where consistent integration of real-world data and mechanistic physiology is essential.

Recent advances in deep generative modeling offer new opportunities to address this gap. Deep generative models provide flexible tools for learning complex, high-dimensional data distributions and synthesizing new samples that follow the same underlying structure (12, 13). By learning the joint distribution of multiple physiological components simultaneously, they can generate virtual subjects whose multivariate profiles reflect realistic developmental relationships rather than treating each component independently. In addition, biological or mechanistic constraints can be incorporated into the training process to ensure that synthesized profiles remain physiologically plausible. Within this framework, latent-space representations emerge as low-dimensional coordinate systems that capture the principal axes of physiological variability and enable smooth interpolation, targeted manipulation, and inversion of generated physiological profiles. Latent representation learning has been increasingly recognized as a unifying concept for modeling high-dimensional biological data, enabling compact, interpretable representations that support generative modeling and translational applications (14).

In this proof-of-concept study, we developed a physiology-informed conditional variational autoencoder (cVAE) that integrates real-world pediatric data with mechanistic model predictions to generate biologically coherent physiological profiles across childhood. By learning a smooth, interpretable latent space conditioned on age and sex, the framework enables physiologically consistent and targeted virtual patient generation and supports PBPK simulations spanning the full pediatric age spectrum.

## METHODS

### Data and Physiological Models

Real-world pediatric data were obtained from the National Health and Nutrition Examination Survey (NHANES) database (9), filtered to include 22,032 individuals from birth to 18 years of age. The dataset included age, sex, height, and weight as core variables, with serum creatinine (SCR) available only for individuals aged 12 years and older. To construct a physiologically coherent dataset, additional variables such as eGFR, organ weights, blood flows, and SCR (for individuals under 12 years) were generated using established mechanistic models tailored to age, sex, and body size (15, 16). For individuals ≥12 years, eGFR was calculated by estimating creatinine production rate (CPR) based on age, sex, and fat-free mass (FFM), and dividing CPR by observed SCR. For those <12 years, SCR was imputed by first simulating physiologically plausible eGFR values with inter-individual variability and then calculating SCR as CPR divided by eGFR. Organ weights were generated using published age- and sex-specific equations (11). Cardiac output (CO) was scaled allometrically by body weight; fat tissue was modeled as a linear function of body weight; other organs were modeled as linear functions of FFM. The complete equations and generated data plots are available in **Supplemental Materials**. The dataset was randomly split into 80% for training (n = 17,625) and 20% for validation (n = 4,407). Data preprocessing and derivation of model-based physiological variables were performed in R version 4.4.0.

### Model Architecture and Training

The cVAE model consists of an encoder, a structured two-dimensional latent space, and a decoder (**Figure 1**). The encoder is a fully connected neural network that receives 32 inputs (30 physiological variables and two conditional inputs) and processes them through two hidden layers with 64 and 32 units using Rectified Linear Unit (ReLU) activation. It outputs four parameters representing the mean (μ) and log-variance, corresponding to the variance (σ²), of a Gaussian posterior. The mean specifies the central location of the latent representation, while the variance reflects its spread, and latent variables are sampled using the reparameterization trick to ensure smooth and continuous interpolation across the latent space. The decoder takes the sampled two-dimensional latent vector concatenated with the conditional inputs (age and sex) and passes them through two hidden layers with 64 and 32 ReLU units, followed by a 30-unit sigmoid output layer. All inputs and outputs were scaled to the [0,1] range using Min–Max normalization.

**Figure 1.**
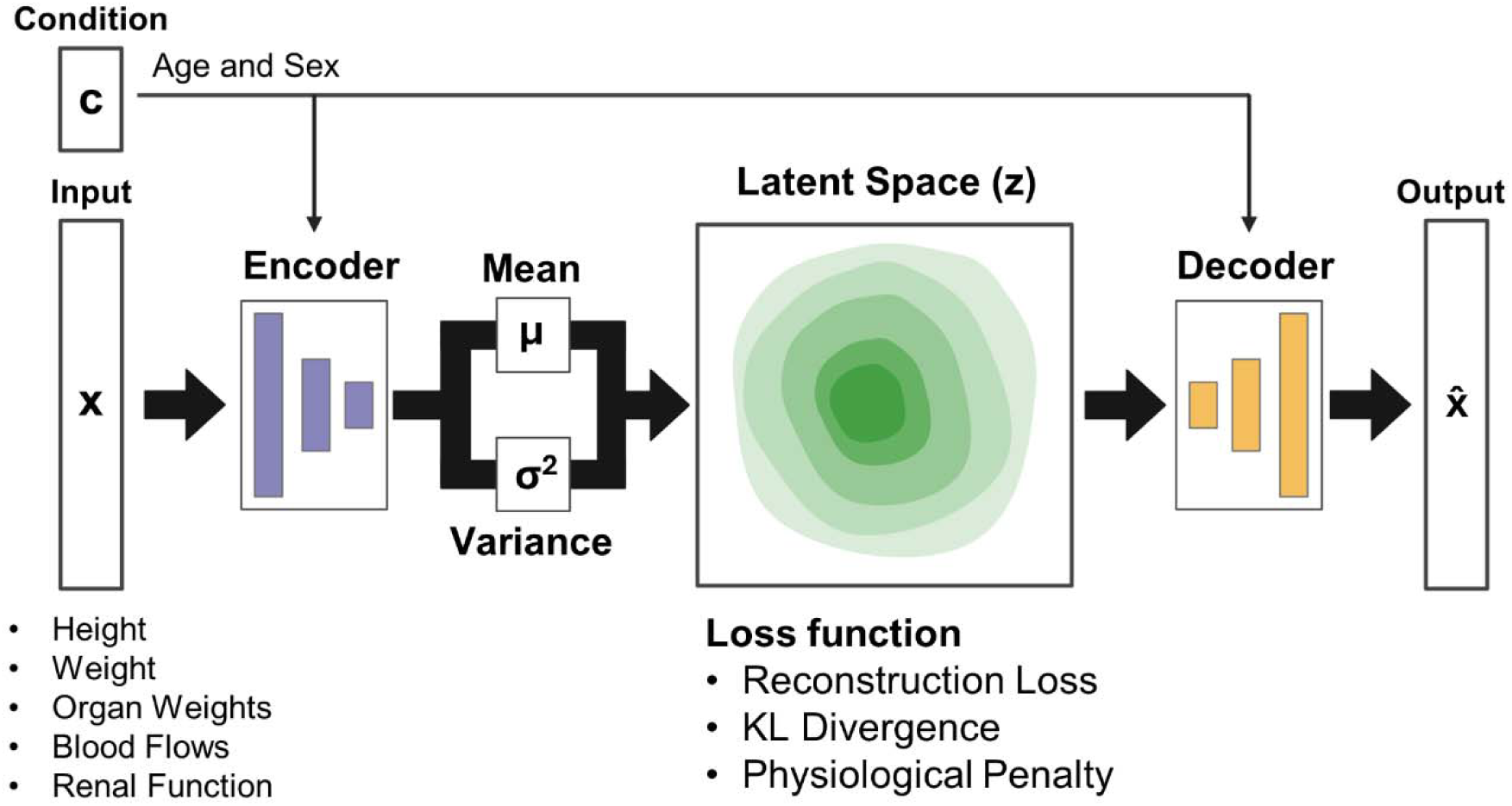
Structure of the physiology informed conditional variational autoencoder (cVAE). The physiology informed cVAE encodes input variables *x* (e.g., height, weight, organ weights) into a latent representation *z*, conditioned on age and sex (*c*). The encoder outputs the mean (μ) and variance (σ²) of a normal distribution, from which *z* is sampled using the reparameterization trick. The decoder then reconstructs the output x from *z* and *c*. The model is trained using a combination of reconstruction loss, KL divergence, and a physiological penalty to ensure that x closely matches *x*, the latent space remains smooth and structured, and the outputs are biologically plausible.

Model training used a composite objective integrating reconstruction accuracy, latent-spac regularization, and physiological plausibility. Reconstruction loss (mean squared error) ensure fidelity to the input data. The Kullback–Leibler (KL) divergence promotes alignment of the latent distribution with a standard normal prior. Physiological penalties are applied when total organ weights exceed body weight or total blood flows exceed cardiac output, based on inverse-transformed outputs in physical units. This physiology-informed design enables the cVAE to generate virtual pediatric patients that maintain realistic multivariate structure while adhering to known developmental physiology. Optimization used the Adaptive Moment Estimation (Adam) algorithm with an 80 % training split. Loss components and weighting procedures are described in the **Supplemental Materials**. Model training and validation were conducted in Python 3.12.7 using TensorFlow 2.19.0.

### Model Validation

Using the validation dataset (20%), model performance was evaluated by assessing reconstruction accuracy and the structure of the latent space. Reconstruction accuracy was quantified using mean absolute error (MAE), coefficient of determination (R²), and Kolmogorov–Smirnov (KS) tests comparing original and generated variable distributions. Latent space regularity and diversity were assessed using KL divergence. To further characterize the latent space, input-to-latent mappings were visually computed by encoding individual physiological profiles and examining their distribution within the two-dimensional space. Additionally, latent space walks were performed by varying either the latent dimensions or age while holding the other inputs constant, enabling evaluation of smooth and physiologically coherent transitions across the whole pediatric age spectrum.

### Applications of Physiology Informed cVAE

To demonstrate the applicability of the physiology-informed cVAE for downstream modeling, the framework was evaluated using targeted physiological profile generation and PBPK-based simulation studies.

For controlled generation of individual physiological profiles for specific clinical scenarios, latent-space inversion was applied. In this procedure, the latent vector *z* was treated as a trainable variable and optimized using gradient-based methods (Adam optimizer, learning rate 0.01, 500 iterations) until the decoder output matched predefined physiological targets, such as body weight or serum creatinine, for a given age and sex. This application demonstrates that the learned latent space supports physiologically consistent and targeted virtual patient generation.

To evaluate the applicability of virtual pediatric patients generated by the cVAE for PBPK model simulations, a vancomycin PBPK model was developed. Vancomycin was selected as an initial testbed because pharmacokinetic data spanning infancy, childhood, and adolescence are widely available in the literature. Vancomycin-specific parameters, including renal clearance and tissue partition coefficient (Kp) scaling factor, were characterized using reported mean concentration–time profiles at ages from 2.6 days to 7.6 years from Schaad et al (17).

To simulate vancomycin pharmacokinetics across the full pediatric age range, latent vectors were sampled from a standard normal distribution (mean 0, SD 1) and assigned age (0–18 years) and sex to span pediatric development. The decoded physiological parameters were used as inputs to the vancomycin PBPK model. Area under the concentration–time curve (AUC) and maximum concentration (C_max_) were then compared with reported pediatric observations across the pediatric spectrum, including special populations such as patients with renal impairment and obesity (17–40). Details of the PBPK model development, implementation, and simulation procedures are provided in the **Supplemental Materials**.

## RESULTS

### Model Training Performance

The cVAE was trained for 300 epochs with a batch size of 64 using a dataset of 17,625 pediatric subjects, combining real-world data and corresponding physiologically modeled variables for each subject. **Figure S1** shows the training dynamics, including the trajectories of the reconstruction loss, KL divergence, and physiological penalty terms (ratios of total organ weights to body weight and total organ blood flows to cardiac output) over the course of training. All components of the loss function stabilized within the first 200 epochs, and no further overfitting was observed beyond that point. The final model achieved a mean absolute error (MAE) of 0.00419 across all variables scaled to the [0, 1] range, with an overall coefficient of determination (R²) of 0.998 when comparing reconstructed outputs to the original data. The final KL divergence between the learned latent distribution and the standard normal prior was 2.11, indicating effective regularization of the latent space.

**Figure S1.**
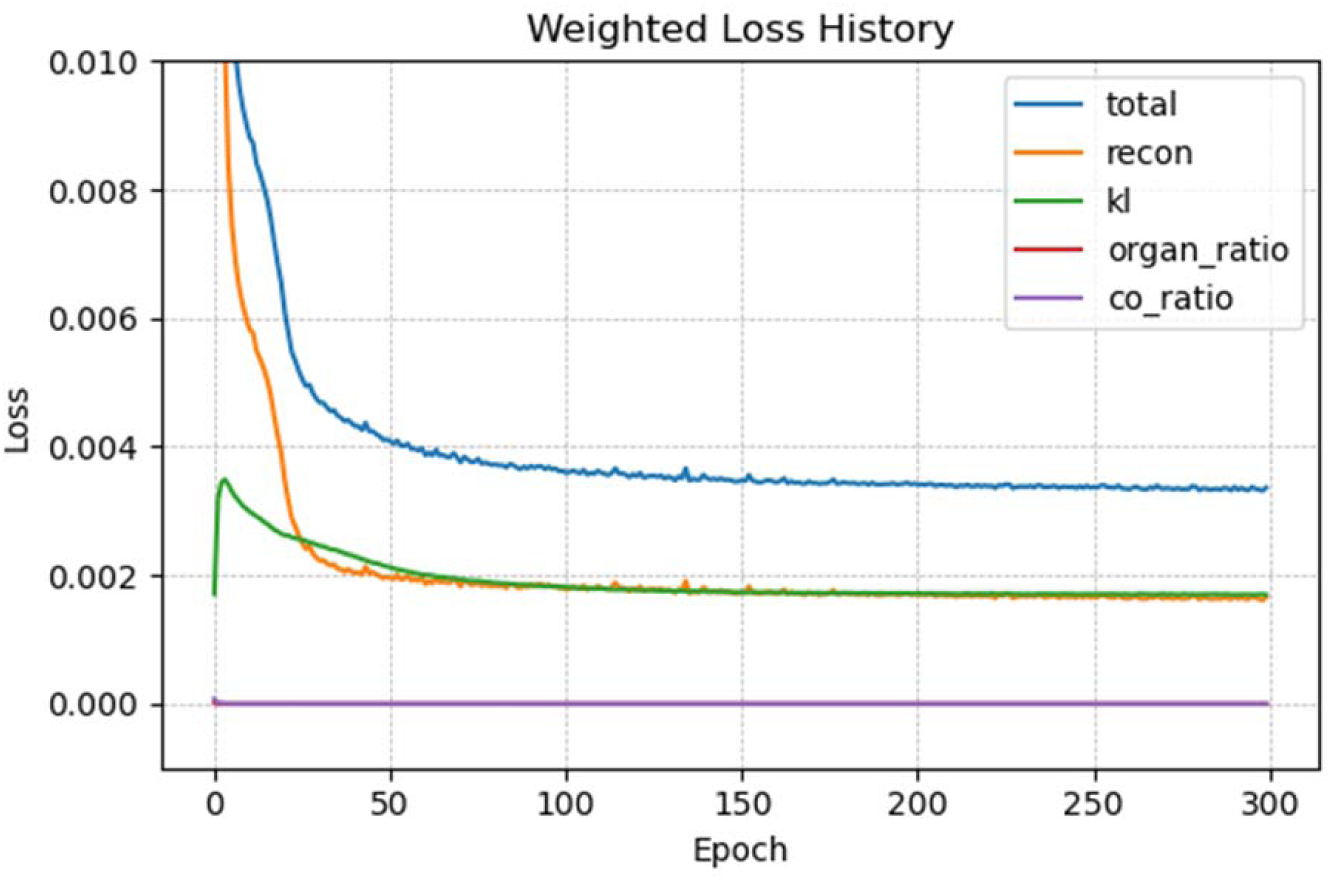
Trajectories of the total loss and its components over training epochs: reconstruction loss (recon), Kullback–Leibler divergence loss (kl), organ weight ratio loss (organ_ratio), and cardiac output ratio loss (co_ratio).

### Validation Accuracy and Latent Space Properties

Quantitative evaluation on the validation set (n = 4,407) confirmed accurate reconstruction of physiological variables. As shown in **Table 1**, the model achieved low MAE and high R² value across all scaled variables. Reconstructed distributions closely matched the original data (**Figure S2**), with non-significant KS test results indicating no major deviations (**Table 1**). Mean validation performance showed an MAE of 0.0043, an R² of 0.998, and a KL divergence of 4.2. To further assess the latent space, input-to-latent mappings were examined by encoding individual physiological profiles and visualizing their distribution within the two-dimensional space. The resulting 3D projections revealed smooth gradients across multiple physiological attributes, suggesting that the latent space is well-structured and biologically meaningful (**Figure 2**). In addition, generated samples were physiologically plausible: all organ weight–to–body weight and organ blood flow–to–cardiac output ratios remained below 1.0 (**Figure 3**), reflecting internal consistency and adherence to structural constraints.

**Table 1.**
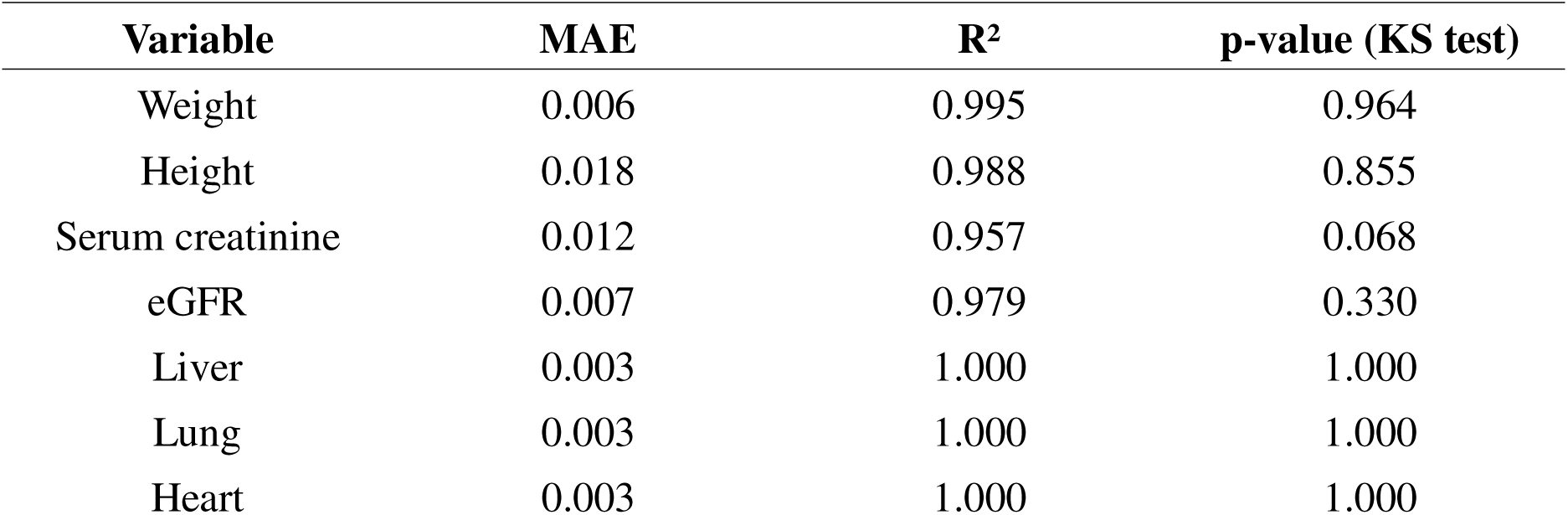
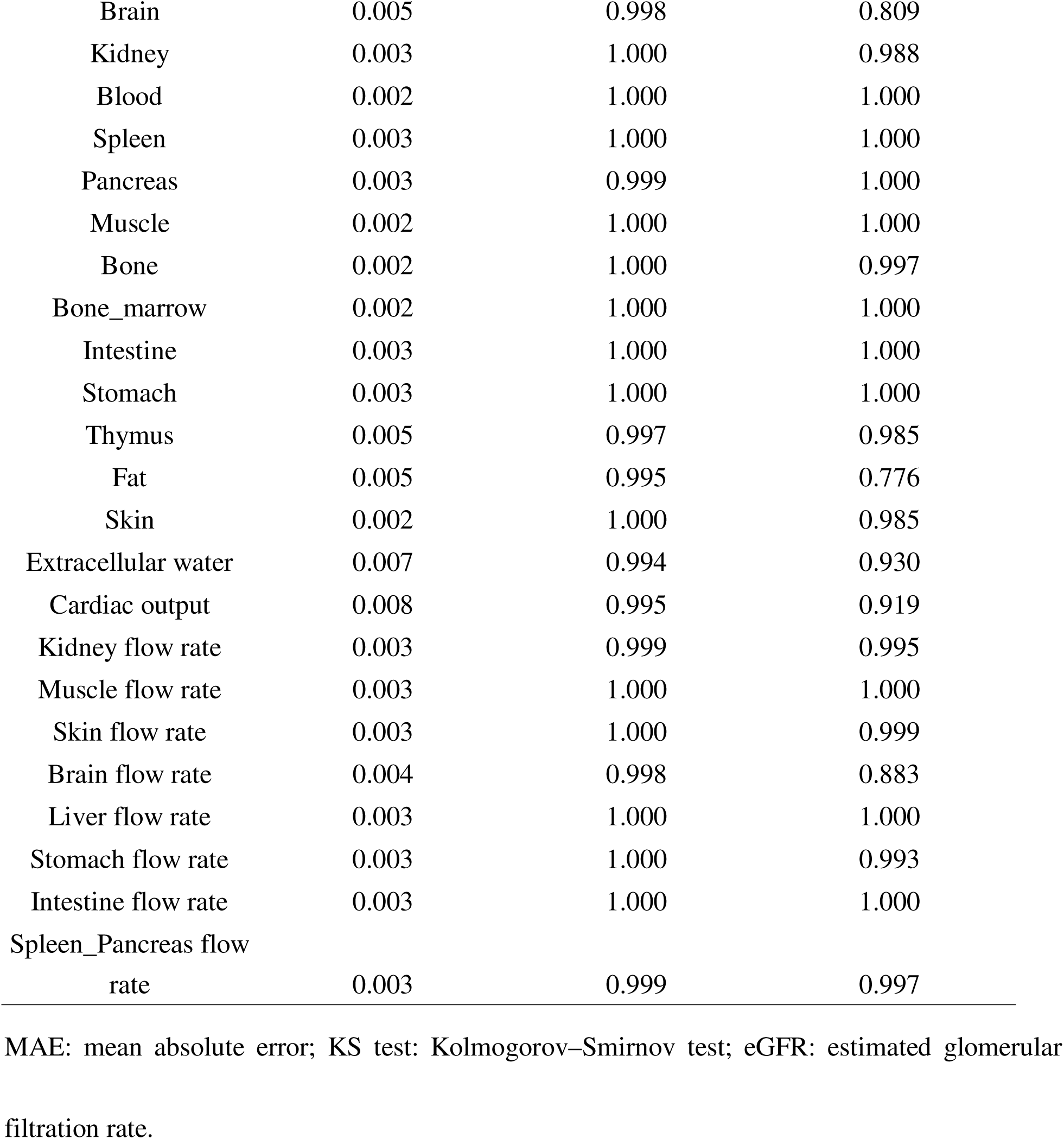
Mean absolute errors (MAE), R² values, and Kolmogorov–Smirnov (KS) test results in the validation dataset.

| Variable | MAE | $R^2$ | p-value (KS test) |
| --- | --- | --- | --- |
| Weight | 0.006 | 0.995 | 0.964 |
| Height | 0.018 | 0.988 | 0.855 |
| Serum creatinine | 0.012 | 0.957 | 0.068 |
| eGFR | 0.007 | 0.979 | 0.330 |
| Liver | 0.003 | 1.000 | 1.000 |
| Lung | 0.003 | 1.000 | 1.000 |
| Heart | 0.003 | 1.000 | 1.000 |
| Brain | 0.005 | 0.998 | 0.809 |
| Kidney | 0.003 | 1.000 | 0.988 |
| Blood | 0.002 | 1.000 | 1.000 |
| Spleen | 0.003 | 1.000 | 1.000 |
| Pancreas | 0.003 | 0.999 | 1.000 |
| Muscle | 0.002 | 1.000 | 1.000 |
| Bone | 0.002 | 1.000 | 0.997 |
| Bone_marrow | 0.002 | 1.000 | 1.000 |
| Intestine | 0.003 | 1.000 | 1.000 |
| Stomach | 0.003 | 1.000 | 1.000 |
| Thymus | 0.005 | 0.997 | 0.985 |
| Fat | 0.005 | 0.995 | 0.776 |
| Skin | 0.002 | 1.000 | 0.985 |
| Extracellular water | 0.007 | 0.994 | 0.930 |
| Cardiac output | 0.008 | 0.995 | 0.919 |
| Kidney flow rate | 0.003 | 0.999 | 0.995 |
| Muscle flow rate | 0.003 | 1.000 | 1.000 |
| Skin flow rate | 0.003 | 1.000 | 0.999 |
| Brain flow rate | 0.004 | 0.998 | 0.883 |
| Liver flow rate | 0.003 | 1.000 | 1.000 |
| Stomach flow rate | 0.003 | 1.000 | 0.993 |
| Intestine flow rate | 0.003 | 1.000 | 1.000 |
| Spleen_Pancreas flow<br>rate | 0.003 | 0.999 | 0.997 |

MAE: mean absolute error; KS test: Kolmogorov–Smirnov test; eGFR: estimated glomerular filtration rate.

**Figure S2.**
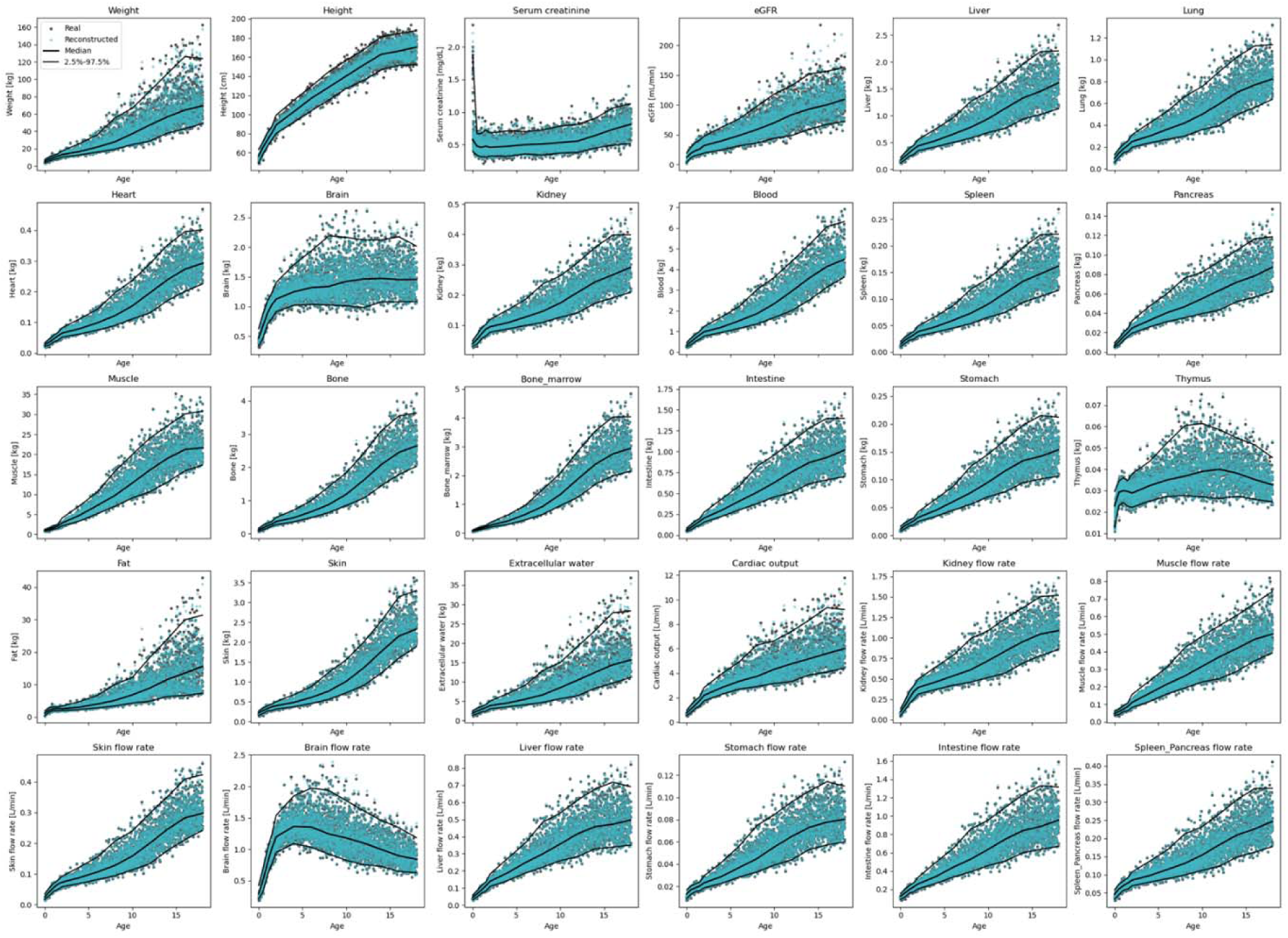
Comparison of reconstructed values (green dots) and original data (gray dots) in the validation dataset. Black lines represent the median and the 2.5th and 97.5th percentiles of the original data.

**Figure 2.**
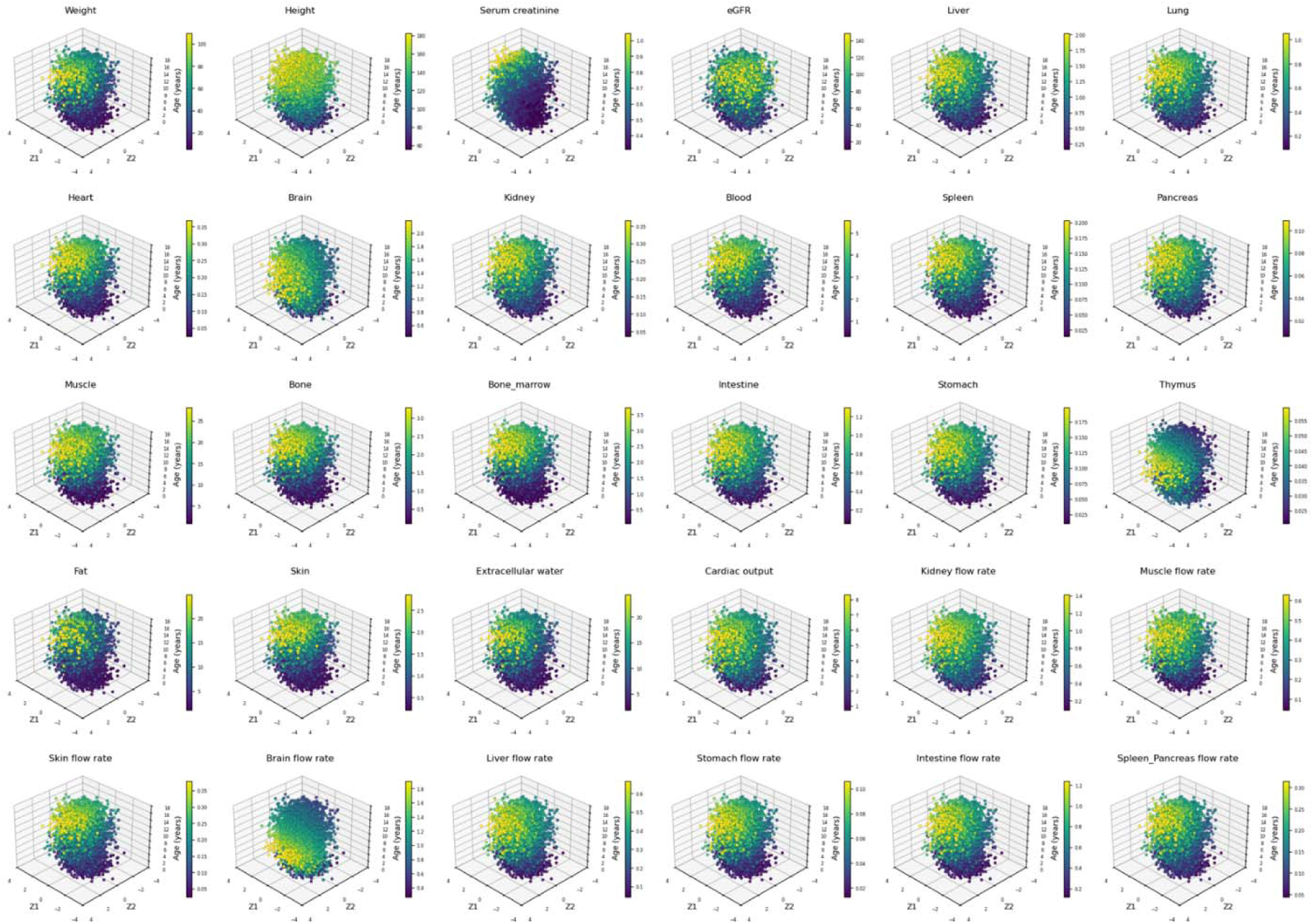
Three-dimensional mapping of the 2D latent space (Z1 and Z2) and age (0–18 years) in the validation dataset. Each point represents an individual subject encoded into the latent space, with the color scale indicating the magnitude of the corresponding physiological variable.

**Figure 3.**
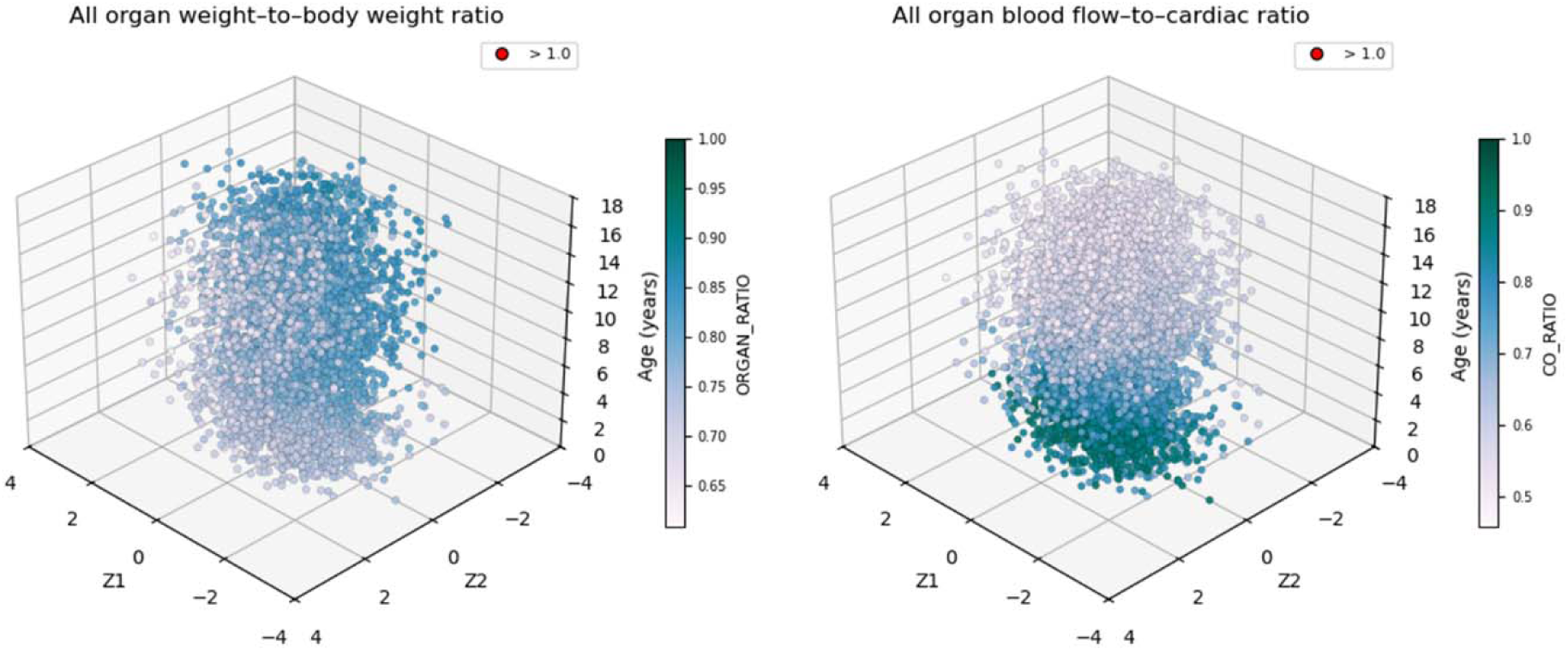
Three-dimensional plots showing the ratios of total organ weights to body weight (left) and total organ blood flows to cardiac output (right) in the validation dataset. No value exceeded 1.0; red dots would indicate such cases if they exist.

Furthermore, when age was varied while keeping latent variables fixed, latent space walks showed that the decoder generated smooth trajectories in outputs over the age range (**Figure 4**), capturing age-dependent physiological maturation. Conversely, when age and sex were fixed (an 8-year-old male), outputs varied smoothly across the latent space (**Figure 5**), indicating locally coherent and interpretable representations.

**Figure 4.**
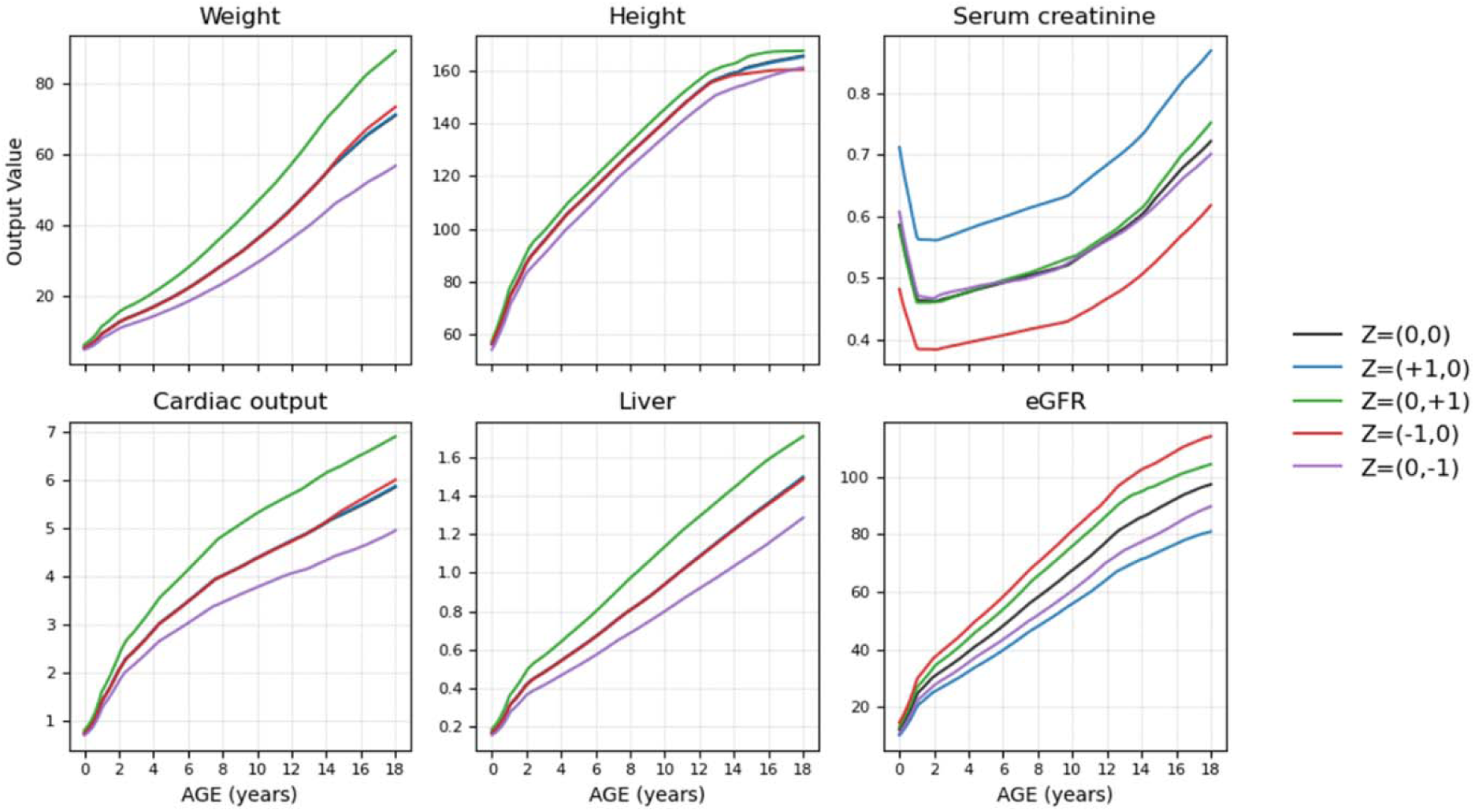
Latent space walk with various Z values across 0-18 age ranges. Physiological outputs were generated by varying age while holding latent variables Z1 and Z2 constant.

**Figure 5.**
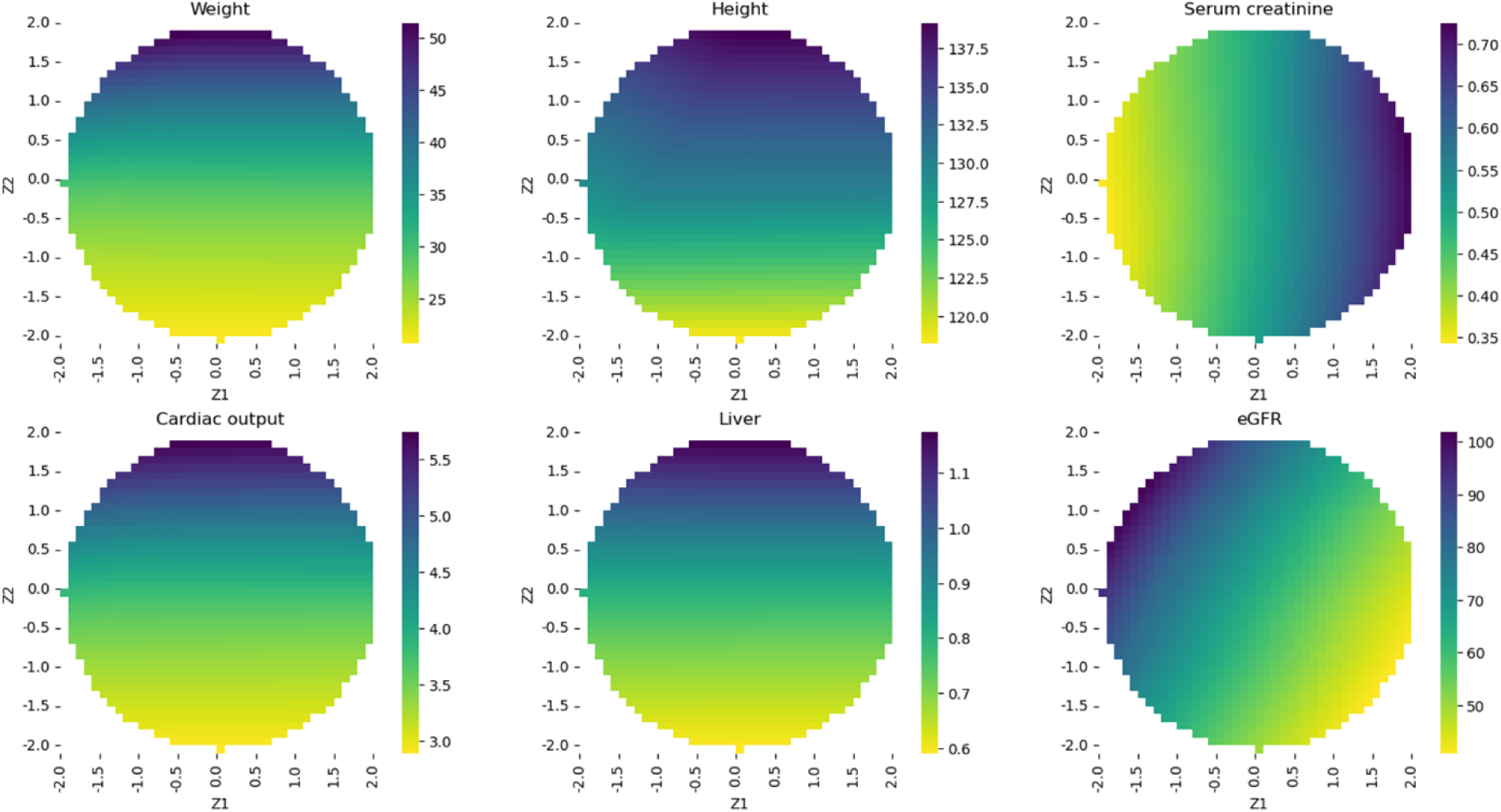
Latent space walk for an 8-year-old male. Physiological outputs were generated by varying latent variables while fixing age and sex, with colors indicating the magnitude of the corresponding physiological variable.

### Latent Space Inversion and Application for PBPK simulations

Latent space inversion was used to generate controlled physiological profiles from patient-specific inputs. For a representative case (an 8-year-old male with a body weight of 35 kg and a serum creatinine level of 0.6 mg/dL), the optimized latent coordinates were estimated as Z1 = 0.920 and Z2 = 0.600 using the Adam optimizer. To introduce controlled variability, Gaussian noise with standard deviations of 0, 0.5, and 1.0 was added to the optimized latent vector prior to decoding. The generated physiological profiles (**Table S1**) remained highly consistent with the target values under low-noise conditions and exhibited physiologically plausible variability at higher noise levels. These results demonstrate that the learned latent space supports accurate, patient-specific profile generation with controllable variability.

In vancomycin PBPK modeling, reported mean concentration–time profiles spanning ages from 2.6 days to 7.6 years from Schaad et al. (17) were well captured using physiological inputs generated by the cVAE, including blood flows and organ weights, matched by age and body weight (**Figure S3**). The estimated mean Kp scaling factor was 3.32 with a coefficient of variation of 18%, and the estimated renal clearance values were strongly correlated with age-specific typical eGFR (R² = 0.9517). Based on these findings, the estimated Kp scaling factor and its variability were applied, and renal clearance was assumed to be fully described by eGFR.

Using this PBPK framework, 500,000 virtual pediatric patients were generated by sampling latent vectors (z1 and z2) from a standard normal distribution (mean 0, SD 1). Vancomycin pharmacokinetics following a 10 mg/kg dose were simulated across the age range of 0 to 18 years. The simulated AUC and C_max_ exhibited smooth age-dependent trends, and both developmental patterns and variability were consistent with reported clinical data (**Table S2** and **Table S3**), including special populations such as patients with renal impairment and obesity (**Figure 6**). These results demonstrate that virtual patients generated by the cVAE can be used to support PBPK modeling across the full pediatric spectrum.

**Figure 6.**
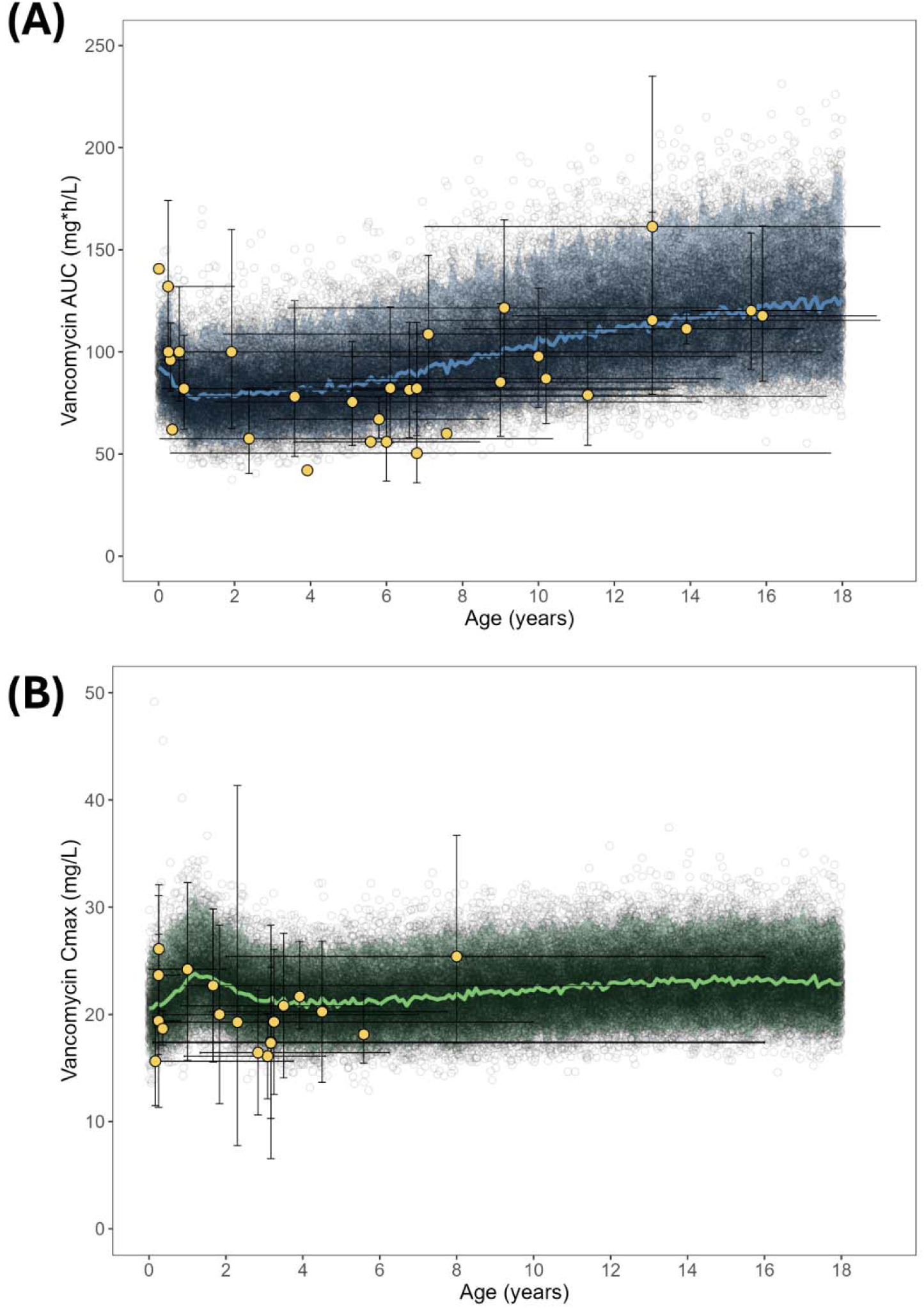
PBPK model-predicted (A) area under the concentration–time curve (AUC) and (B) maximum concentration (Cmax) of vancomycin in pediatric patients aged 0–18 years. Open black circles represent PBPK model predictions, and the shaded area indicates the 95% prediction interval. Yellow dots denote reported observations, with horizontal and vertical error bars representing the reported age range and AUC or Cmax variability (range, interquartile range, or standard deviation, when available).

**Table S1.** Generated data using latent space inversion for an 8-year-old male (35 kg, serum creatinine 0.6 mg/dL), decoded with Gaussian noise (SD = 0, 0.5, and 1.0).

| Noise | ID | AGE | SEX | Weight | Height | eGFR | Serum creatinine | eGFR | Cardiac output | Liver |
| --- | --- | --- | --- | --- | --- | --- | --- | --- | --- | --- |
| 0 | 0 | 8 | 0 | 35.00 | 132.7 | 57.86 | 0.60 | 57.86 | 4.30 | 0.90 |
|  | 1 | 8 | 0 | 35.00 | 132.7 | 57.86 | 0.60 | 57.86 | 4.30 | 0.90 |
|  | 2 | 8 | 0 | 35.00 | 132.7 | 57.86 | 0.60 | 57.86 | 4.30 | 0.90 |
|  | 3 | 8 | 0 | 35.00 | 132.7 | 57.86 | 0.60 | 57.86 | 4.30 | 0.90 |
|  | 4 | 8 | 0 | 35.00 | 132.7 | 57.86 | 0.60 | 57.86 | 4.30 | 0.90 |
|  | 5 | 8 | 0 | 35.00 | 132.7 | 57.86 | 0.60 | 57.86 | 4.30 | 0.90 |
|  | 6 | 8 | 0 | 35.00 | 132.7 | 57.86 | 0.60 | 57.86 | 4.30 | 0.90 |
|  | 7 | 8 | 0 | 35.00 | 132.7 | 57.86 | 0.60 | 57.86 | 4.30 | 0.90 |
|  | 8 | 8 | 0 | 35.00 | 132.7 | 57.86 | 0.60 | 57.86 | 4.30 | 0.90 |
|  | 9 | 8 | 0 | 35.00 | 132.7 | 57.86 | 0.60 | 57.86 | 4.30 | 0.90 |
| Noise | ID | AGE | SEX | Weight | Height | eGFR | Serum creatinine | eGFR | Cardiac output | Liver |
| 0.5 | 0 | 8 | 0 | 40.49 | 135.05 | 68.48 | 0.54 | 68.48 | 4.8 | 1.00 |
|  | 1 | 8 | 0 | 28.53 | 129.86 | 51.08 | 0.61 | 51.08 | 3.69 | 0.78 |
|  | 2 | 8 | 0 | 44.49 | 136.62 | 68.92 | 0.57 | 68.92 | 5.17 | 1.07 |
|  | 3 | 8 | 0 | 32.96 | 131.9 | 70.74 | 0.47 | 70.74 | 4.12 | 0.87 |
|  | 4 | 8 | 0 | 31.12 | 131.1 | 48.56 | 0.67 | 48.56 | 3.94 | 0.83 |
|  | 5 | 8 | 0 | 34.54 | 132.52 | 61.33 | 0.56 | 61.33 | 4.26 | 0.89 |
|  | 6 | 8 | 0 | 32.12 | 131.53 | 48.23 | 0.69 | 48.23 | 4.04 | 0.85 |

|  | 7 | 8 | 0 | 32.98 | 131.9 | 58.74 | 0.57 | 58.74 | 4.12 | 0.87 |
| --- | --- | --- | --- | --- | --- | --- | --- | --- | --- | --- |
|  | 8 | 8 | 0 | 48.00 | 138.14 | 54.54 | 0.75 | 54.54 | 5.46 | 1.13 |
|  | 9 | 8 | 0 | 37.01 | 133.64 | 53.79 | 0.67 | 53.79 | 4.49 | 0.94 |
| Noise | ID | AGE | SEX | Weight | Height | eGFR | Serum creatinine | eGFR | Cardiac output | Liver |
| 1.0 | 0 | 8 | 0 | 46.84 | 137.5 | 81.93 | 0.48 | 81.93 | 5.37 | 1.11 |
|  | 1 | 8 | 0 | 24.77 | 125.11 | 45.75 | 0.63 | 45.75 | 3.31 | 0.69 |
|  | 2 | 8 | 0 | 56.56 | 141.46 | 80.53 | 0.53 | 80.53 | 6.15 | 1.26 |
|  | 3 | 8 | 0 | 31.16 | 130.35 | 86.27 | 0.38 | 86.27 | 3.95 | 0.83 |
|  | 4 | 8 | 0 | 27.81 | 129.11 | 40.52 | 0.76 | 40.52 | 3.61 | 0.76 |
|  | 5 | 8 | 0 | 34.08 | 132.34 | 64.98 | 0.52 | 64.98 | 4.22 | 0.89 |
|  | 6 | 8 | 0 | 29.42 | 130.27 | 39.93 | 0.79 | 39.93 | 3.77 | 0.80 |
|  | 7 | 8 | 0 | 31.06 | 131.1 | 59.63 | 0.55 | 59.63 | 3.94 | 0.83 |
|  | 8 | 8 | 0 | 64.94 | 144.56 | 50.27 | 0.90 | 50.27 | 6.91 | 1.38 |
|  | 9 | 8 | 0 | 39.12 | 134.58 | 49.98 | 0.74 | 49.98 | 4.68 | 0.98 |

**Figure S3.**
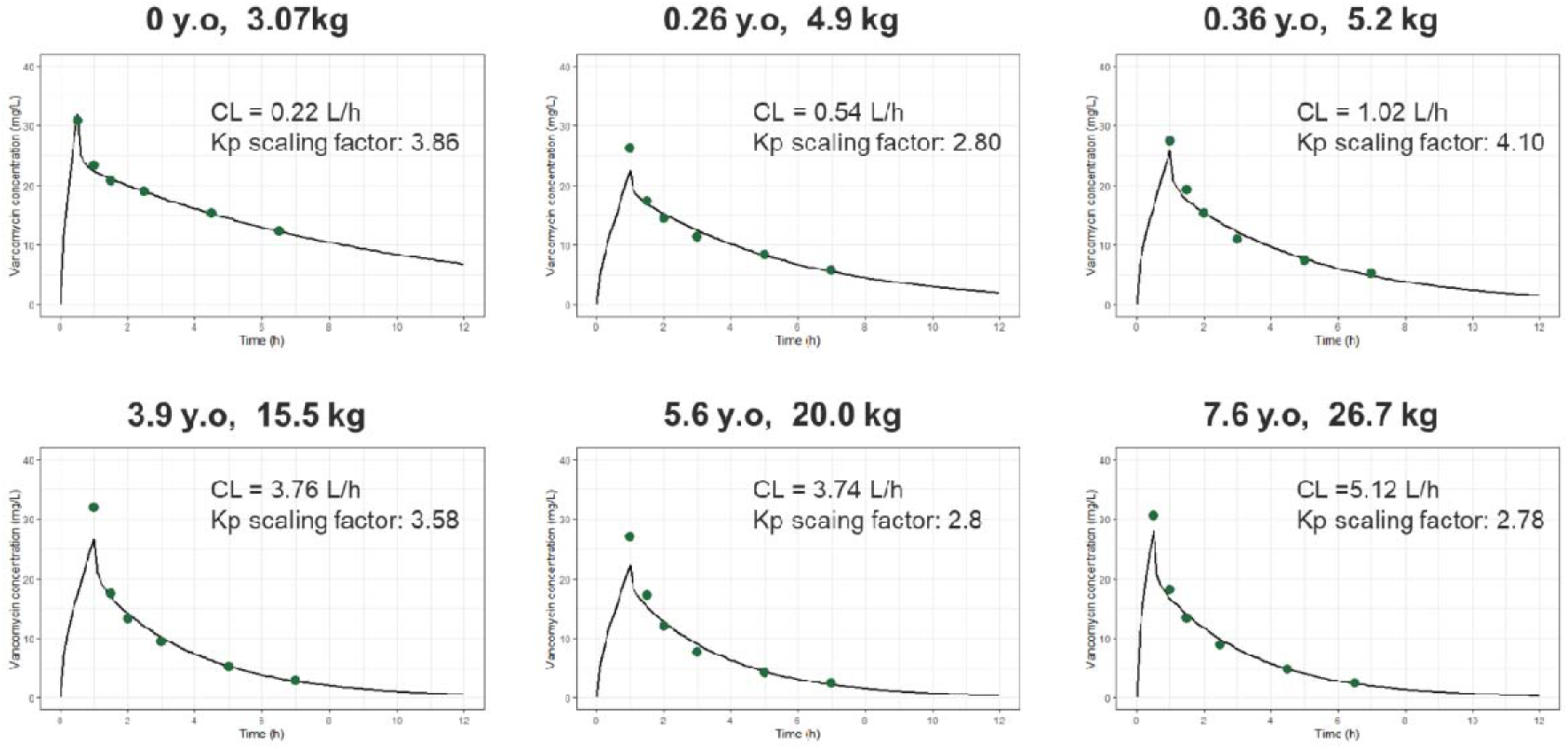
Predicted vancomycin concentration–time profiles from the PBPK model in pediatric patients aged 2.6 days to 7.6 years, based on data reported by Schaad et al (17). Dots represent observed concentrations, and solid lines represent PBPK model–predicted profiles.

**Table S2.**
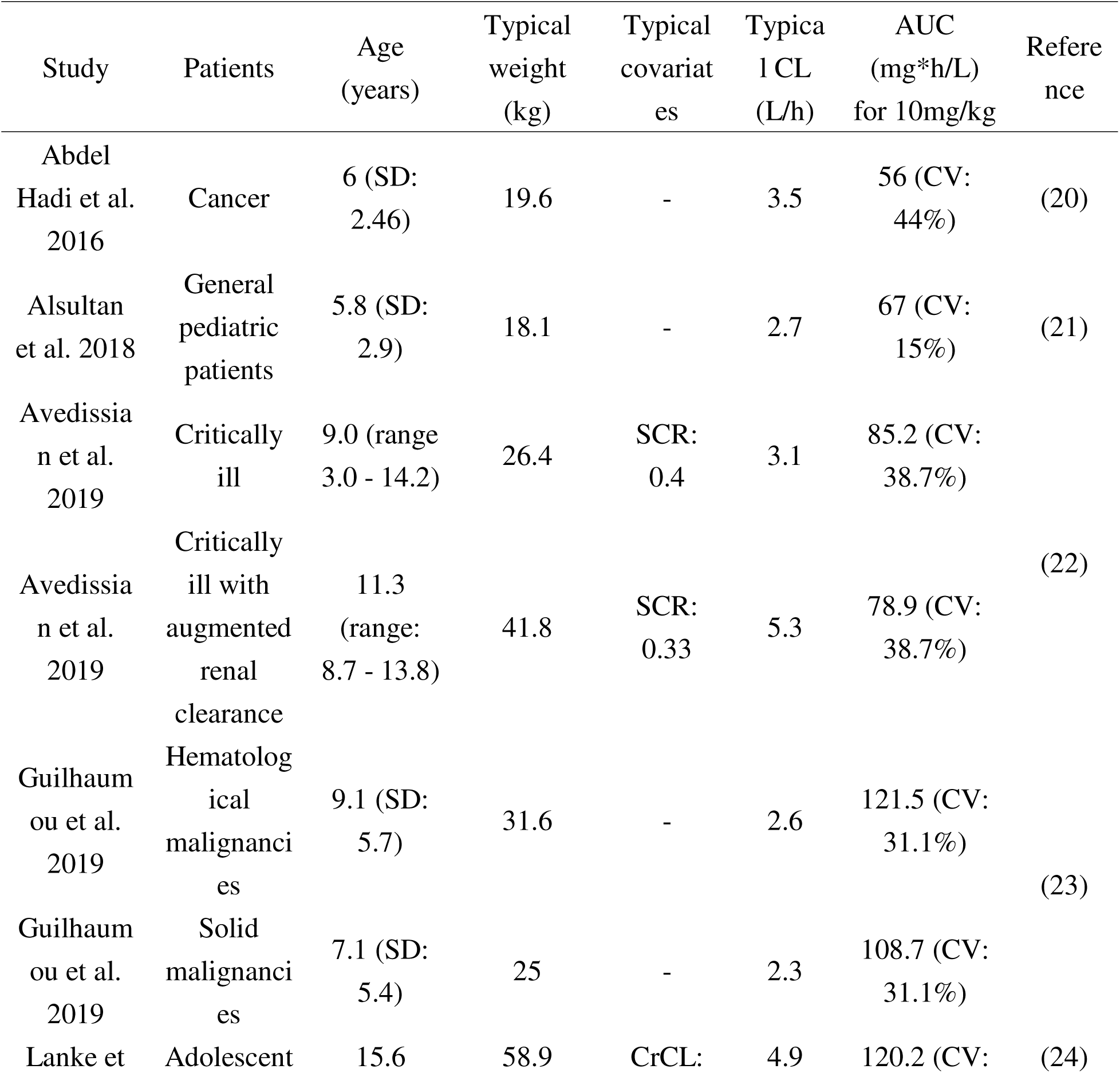
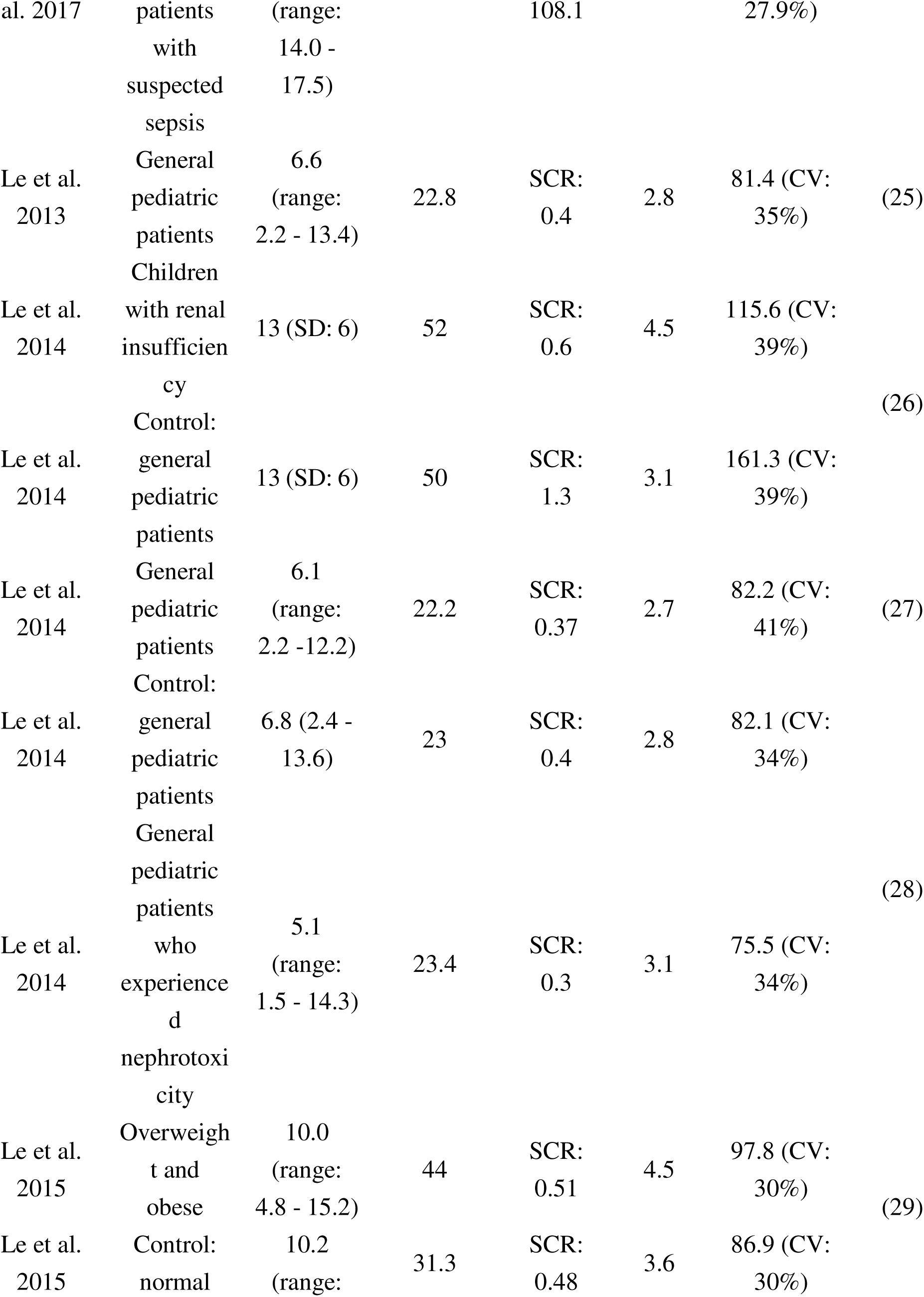
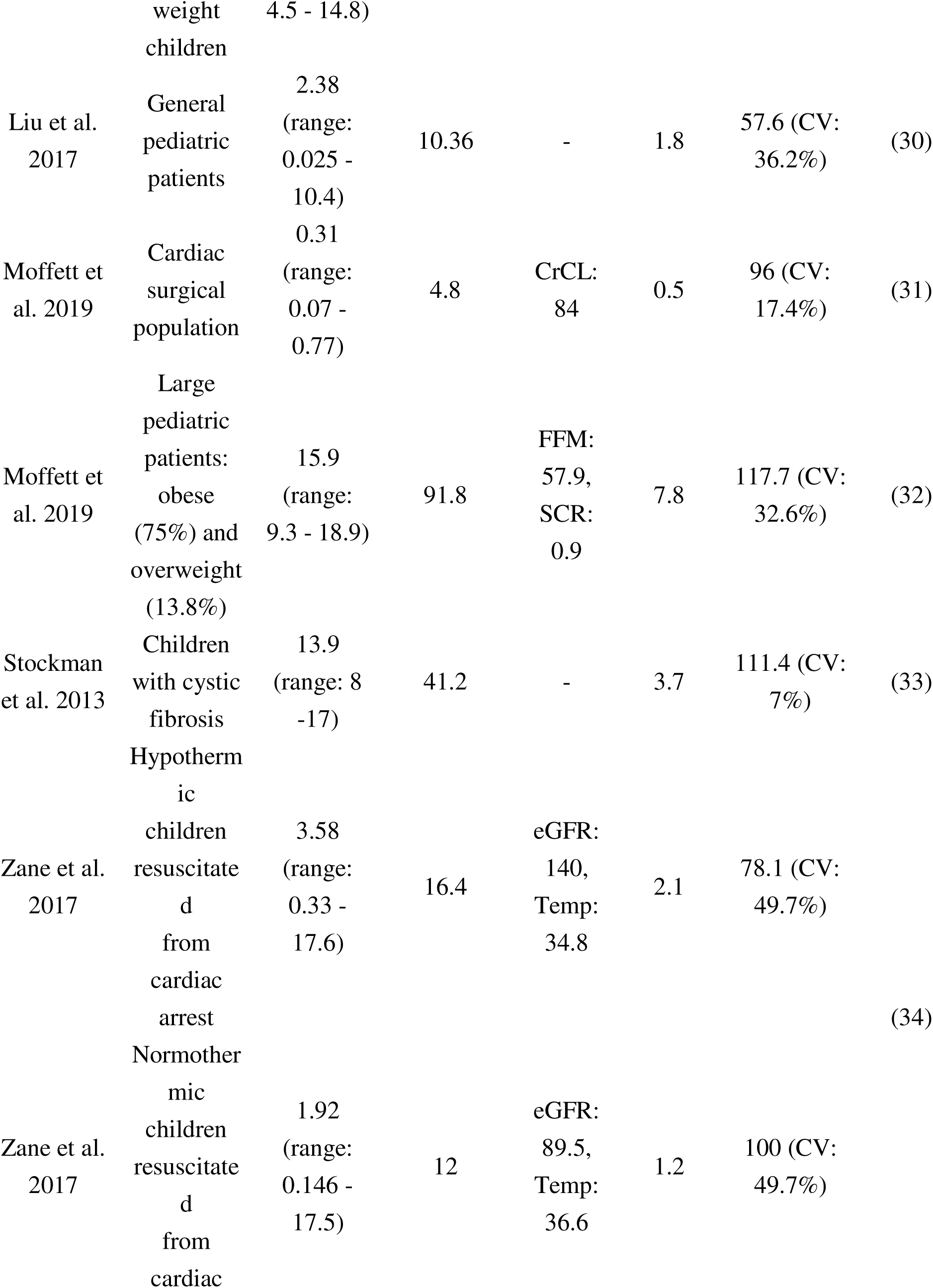
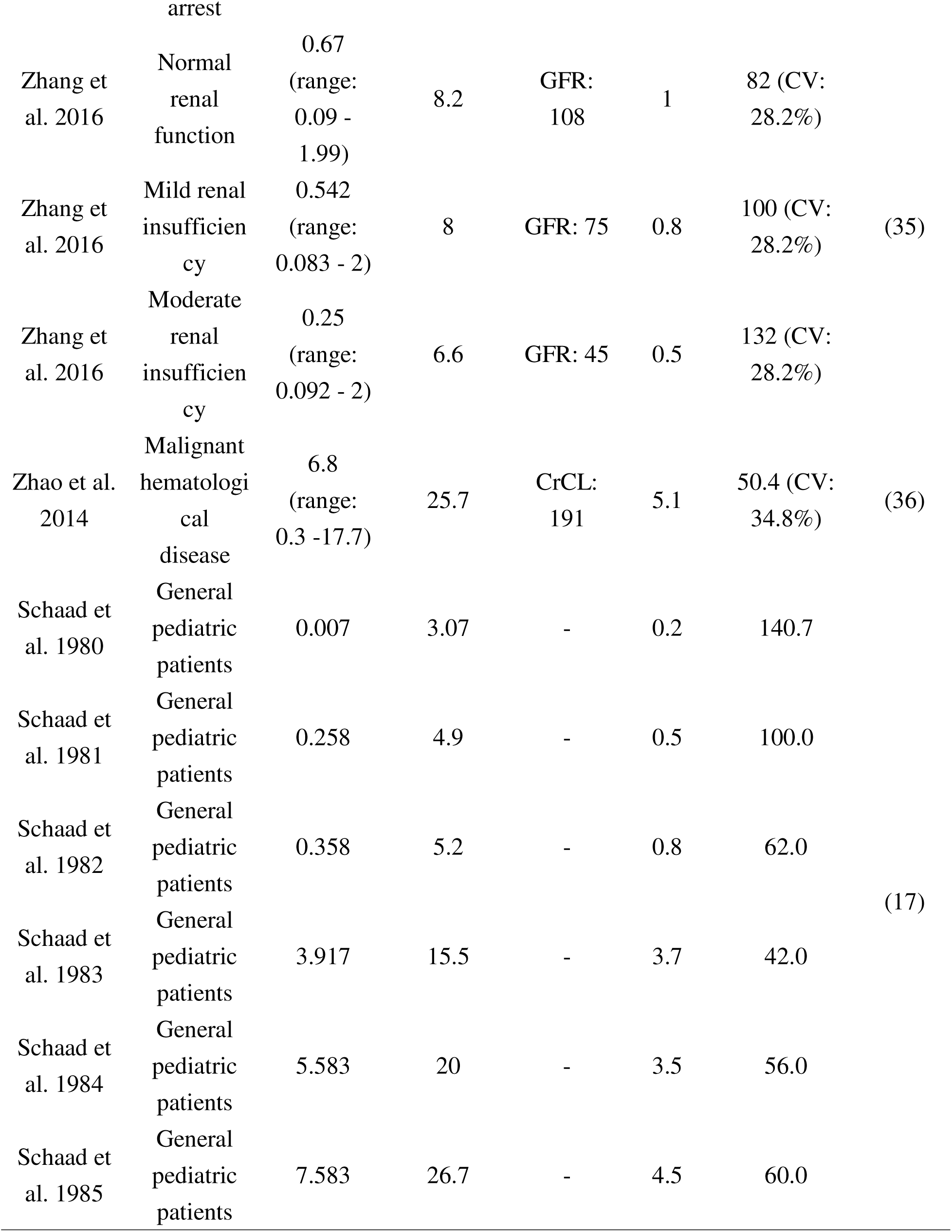
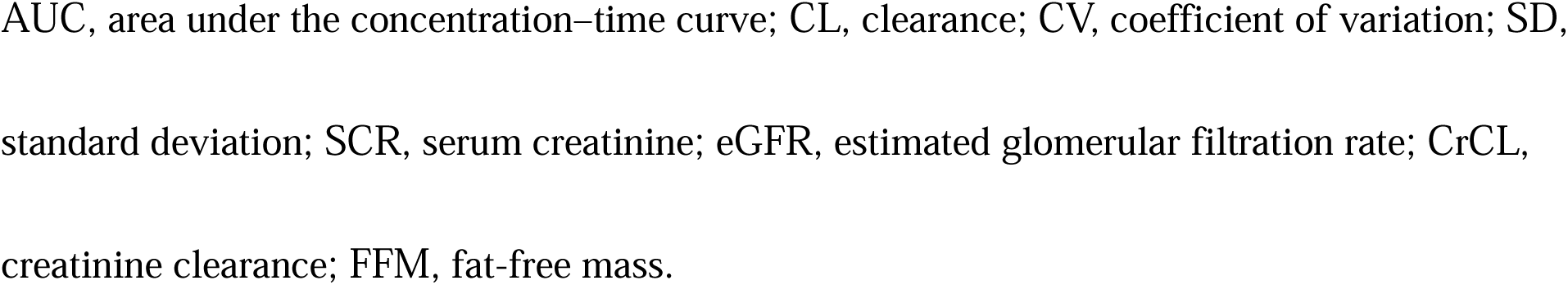
Summary of vancomycin pharmacokinetic studies used for AUC observations.

**Table S3.**
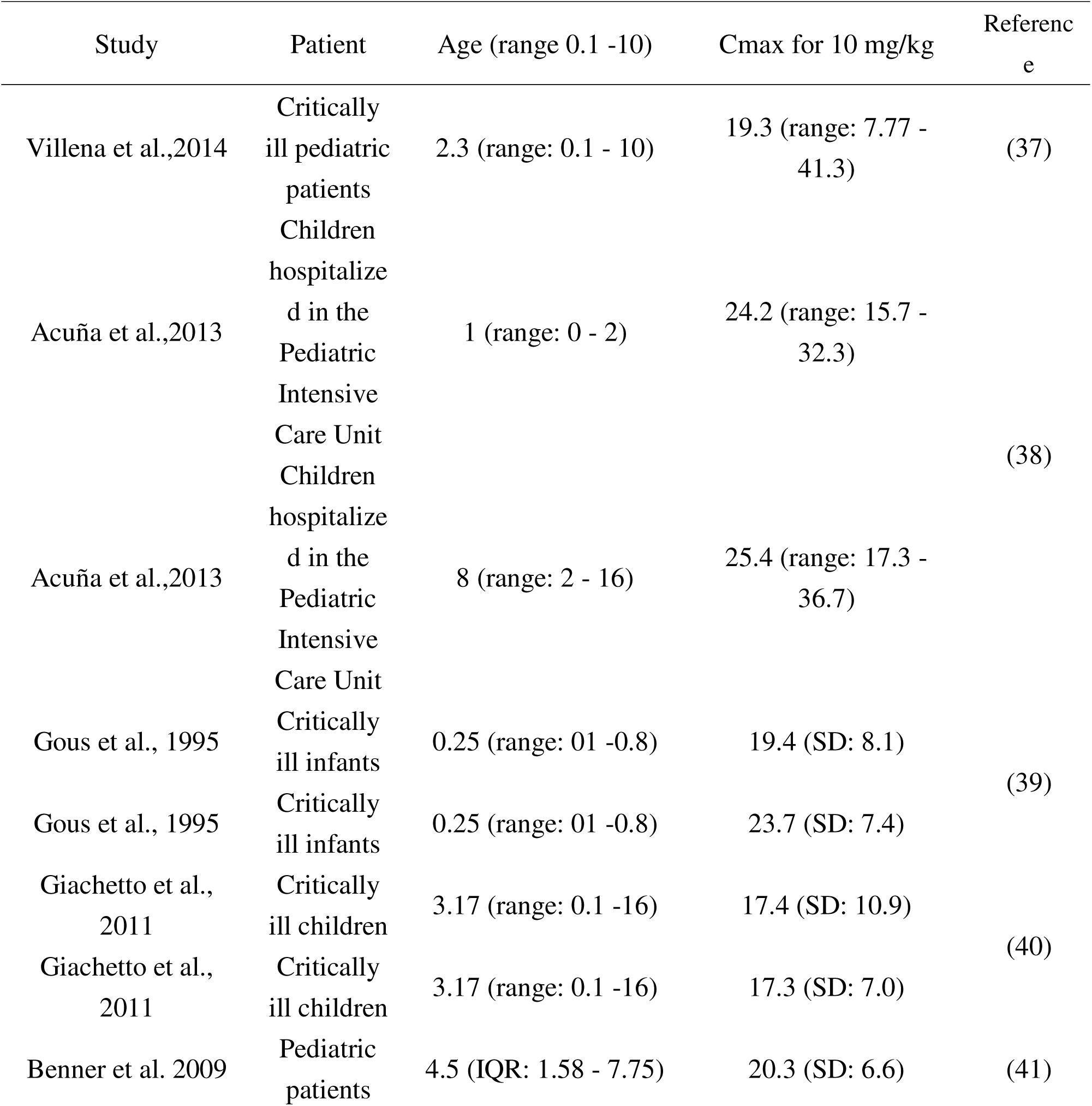
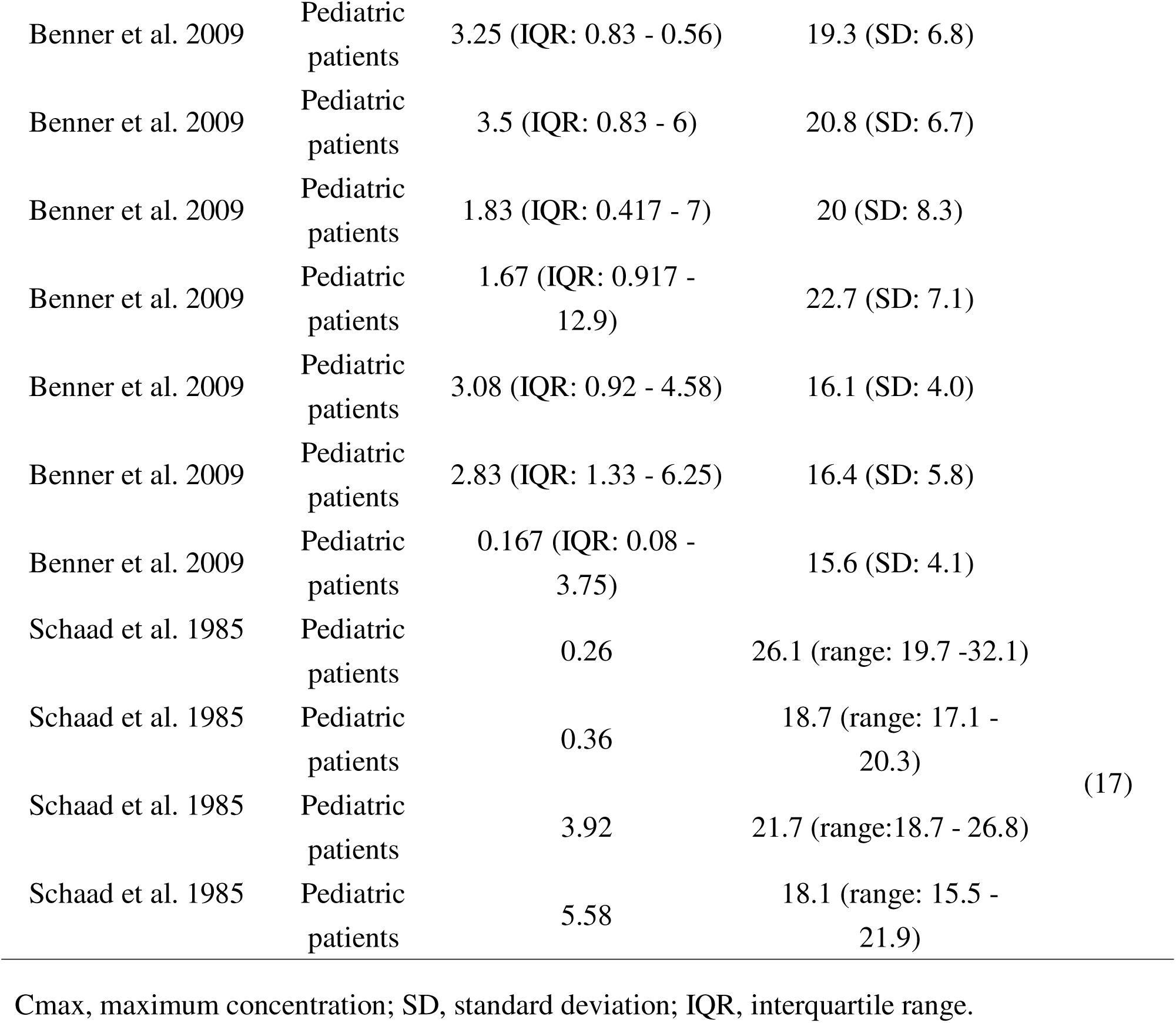
Summary of vancomycin pharmacokinetic studies used for Cmax observations.

## DISCUSSION

This study introduces a physiology-informed cVAE trained on real-world pediatric data and mechanistically derived physiological datasets to generate pediatric virtual patients. The model accurately reconstructs key physiological variables while preserving biological plausibility, and learns a smooth, interpretable latent space that reflects age, sex, and developmental changes. The framework was demonstrated using latent space inversion and vancomycin PBPK modeling, enabling targeted virtual patient generation and PBPK simulations across the full pediatric age range. The resulting multivariate physiological profiles provide coherent and biologically consistent inputs for pediatric model-informed simulations.

Recent advances in deep generative modeling, such as generative adversarial networks (GANs) and VAEs, offer new solutions for clinical data generation. While GANs produce realistic samples, they often suffer from limited stability and interpretability. VAEs provide a structured latent space, supporting stable and diverse generation (12, 13). cVAEs further allow controllable sampling and have been applied in biomedical domains (42–44), offering advantages in controllable sampling, modeling high-dimensional distributions, and generating diverse and realistic outputs. In parallel, physics-informed neural networks (PINNs) (45, 46) embed physical or physiological constraints directly into model training. In this study, we introduced a novel framework that integrates cVAE-based generation with physiology informed loss constraints, enabling the creation of pediatric physiology data that are diverse, age-appropriate, and physiologically coherent. To our knowledge, this represents the first application of such an integrated framework for generating pediatric virtual patients.

The validation results demonstrated that the cVAE achieves both high reconstruction accuracy and a well-structured latent space. Because reconstruction loss and KL divergence often involve a trade-off in variational autoencoders (47), the model was carefully tuned to reproduce the original data distribution accurately while keeping the KL divergence low. In the validation set, the KL divergence was approximately twice that of the training set, which is a typical pattern in VAEs when applied to unseen data and considered acceptable (48). The latent space remained smooth and well-organized across the validation set as well, as demonstrated by the latent space walk. These findings support the interpretation that the learned latent space represents physiologically meaningful structure across pediatric development.

Through latent-space inversion, the latent space enables the generation of targeted physiological profiles corresponding to specific clinical scenarios by optimizing latent variables to match predefined characteristics. Beyond targeted generation, the latent space can also be broadly sampled across the pediatric age range to create diverse virtual patient populations. These physiologically coherent profiles were subsequently integrated into a PBPK model to predict vancomycin pharmacokinetics across childhood. The smooth age-dependent behavior observed in the PBPK simulations reflects the continuity of physiological maturation encoded within the latent space. In addition, the ability to accommodate special populations, such as individuals with renal impairment or obesity, indicates that the generated physiological profiles retain sufficient flexibility to support heterogeneous clinical scenarios. Collectively, these results suggest that integrating physiology-informed generative models with PBPK frameworks provides a scalable approach for constructing pediatric in silico populations, with potential applications in virtual clinical trials and digital twin development.

Several limitations merit attention. The current model successfully generates physiologically plausible profiles but does not yet incorporate all variables critical to pediatric applications, such as enzyme and transporter expression levels and their ontogeny, which are essential for predicting drug disposition with PBPK modeling in pediatric populations (49). Additionally, incorporating more granular physiological constraints, rather than relying only on limits for total organ weights or total blood flows, may be warranted to further enhance the biological realism of generated data. Incorporating these features will be crucial for future model refinement.

In conclusion, we established a physiology-informed cVAE that integrates real-world pediatric data with mechanistic physiology to generate biologically coherent pediatric virtual patients. The model reconstructs key variables with biological plausibility and yields an interpretable latent space that encodes age, sex, and developmental trajectories of physiological parameters. The framework was applied to latent space inversion and vancomycin PBPK modeling to enable targeted virtual patient generation and PBPK simulations across the full pediatric age range. Continued integration of clinical data and biological details will strengthen the framework to support both precision medicine and drug development in pediatric populations.

## Data Availability

All data produced in the present study are available upon reasonable request to the authors

## Ethics Approval

Not applicable

## Author Contributions

K.I. and T.M. designed the study, performed the analysis, and wrote the manuscript.

## Acknowledgements

The authors used ChatGPT (OpenAI) to improve the readability, clarity, and language of this manuscript. The authors reviewed and edited the output and take full responsibility for the content, interpretation, and conclusions of the publication.

## REFERENCES

(1) Domingues, C., Jarak, I., Veiga, F., Dourado, M. & Figueiras, A. Pediatric Drug Development: Reviewing Challenges and Opportunities by Tracking Innovative Therapies. Pharmaceutics 15, (2023).

(2) Kearns, G.L., Abdel-Rahman, S.M., Alander, S.W., Blowey, D.L., Leeder, J.S. & Kauffman, R.E. Developmental pharmacology--drug disposition, action, and therapy in infants and children. N Engl J Med 349, 1157–67 (2003).

(3) Yellepeddi, V., Rower, J., Liu, X., Kumar, S., Rashid, J. & Sherwin, C.M.T. State-of-the-Art Review on Physiologically Based Pharmacokinetic Modeling in Pediatric Drug Development. Clin Pharmacokinet 58, 1–13 (2019).

(4) Yates, J.W.T. et al. Physiologically-Based Pharmacokinetic Modeling to Support Pediatric Clinical Development: An IQ Working Group Perspective on the Current Status and Challenges. CPT Pharmacometrics Syst Pharmacol, (2025).

(5) Tanaka, R., Irie, K. & Mizuno, T. Physiologically Based Pharmacokinetic Modeling of Antibiotics in Children: Perspectives on Model-Informed Precision Dosing. Antibiotics (Basel) 14, (2025).

(6) van der Heijden, J.E.M., Freriksen, J.J.M., de Hoop-Sommen, M.A., Greupink, R. & de Wildt, S.N. Physiologically-Based Pharmacokinetic Modeling for Drug Dosing in Pediatric Patients: A Tutorial for a Pragmatic Approach in Clinical Care. Clin Pharmacol Ther 114, 960–71 (2023).

(7) Templeton, I.E., Jones, N.S. & Musib, L. Pediatric Dose Selection and Utility of PBPK in Determining Dose. AAPS J 20, 31 (2018).

(8) T’Jollyn, H., Vermeulen, A. & Van Bocxlaer, J. PBPK and its Virtual Populations: the Impact of Physiology on Pediatric Pharmacokinetic Predictions of Tramadol. AAPS J 21, 8 (2018).

(9) Centers for Disease Control and Prevention (CDC), National Health and Nutrition Examination Survey (NHANES). <https://wwwn.cdc.gov/nchs/nhanes/>.

(10) Maharaj, A.R. & Edginton, A.N. Physiologically based pharmacokinetic modeling and simulation in pediatric drug development. CPT Pharmacometrics Syst Pharmacol 3, e150 (2014).

(11) Chang, H.P., Kim, S.J., Wu, D., Shah, K. & Shah, D.K. Age-Related Changes in Pediatric Physiology: Quantitative Analysis of Organ Weights and Blood Flows : Age-Related Changes in Pediatric Physiology. AAPS J 23, 50 (2021).

(12) Gangwal, A. et al. Generative artificial intelligence in drug discovery: basic framework, recent advances, challenges, and opportunities. Front Pharmacol 15, 1331062 (2024).

(13) Mishra, A., Majumder, A., Kommineni, D., Anna Joseph, C., Chowdhury, T. & Anumula, S.K. Role of Generative Artificial Intelligence in Personalized Medicine: A Systematic Review. Cureus 17, e82310 (2025).

(14) Kopf, A. & Claassen, M. Latent representation learning in biology and translational medicine. Patterns (N Y) 2, 100198 (2021).

(15) O’Hanlon, C.J., Holford, N., Sumpter, A. & Al-Sallami, H.S. Consistent methods for fat-free mass, creatinine clearance, and glomerular filtration rate to describe renal function from neonates to adults. CPT Pharmacometrics Syst Pharmacol 12, 401–12 (2023).

(16) Correction to: Consistent methods for fat free mass, creatinine clearance, and glomerular filtration rate to describe renal function from neonates to adults. CPT Pharmacometrics Syst Pharmacol 13, 181-2 (2024).

(17) Schaad, U.B., McCracken, G.H., Jr. & Nelson, J.D. Clinical pharmacology and efficacy of vancomycin in pediatric patients. J Pediatr 96, 119–26 (1980).

(18) Aljutayli, A., El-Haffaf, I., Marsot, A. & Nekka, F. An Update on Population Pharmacokinetic Analyses of Vancomycin, Part II: In Pediatric Patients. Clin Pharmacokinet 61, 47-70 (2022).

(19) Akunne, O.O., Mugabo, P. & Argent, A.C. Pharmacokinetics of Vancomycin in Critically Ill Children: A Systematic Review. Eur J Drug Metab Pharmacokinet 47, 31–48 (2022).

(20) Abdel Hadi, O., Al Omar, S., Nazer, L.H., Mubarak, S. & Le, J. Vancomycin pharmacokinetics and predicted dosage requirements in pediatric cancer patients. J Oncol Pharm Pract 22, 448–53 (2016).

(21) Alsultan, A. et al. Optimizing Vancomycin Monitoring in Pediatric Patients. Pediatr Infect Dis J 37, 880–5 (2018).

(22) Avedissian, S.N. et al. Augmented Renal Clearance Using Population-Based Pharmacokinetic Modeling in Critically Ill Pediatric Patients. Pediatr Crit Care Med 18, e388–e94 (2017).

(23) Guilhaumou, R. et al. Pediatric Patients With Solid or Hematological Tumor Disease: Vancomycin Population Pharmacokinetics and Dosage Optimization. Ther Drug Monit 38, 559–66 (2016).

(24) Lanke, S., Yu, T., Rower, J.E., Balch, A.H., Korgenski, E.K. & Sherwin, C.M. AUC-Guided Vancomycin Dosing in Adolescent Patients With Suspected Sepsis. J Clin Pharmacol 57, 77–84 (2017).

(25) Le, J. et al. Improved vancomycin dosing in children using area under the curve exposure. Pediatr Infect Dis J 32, e155–63 (2013).

(26) Le, J. et al. Population-Based Pharmacokinetic Modeling of Vancomycin in Children with Renal Insufficiency. J Pharmacol Clin Toxicol 2, 1017–26 (2014).

(27) Le, J. et al. Vancomycin monitoring in children using bayesian estimation. Ther Drug Monit 36, 510–8 (2014).

(28) Le, J. et al. Pharmacodynamic Characteristics of Nephrotoxicity Associated With Vancomycin Use in Children. J Pediatric Infect Dis Soc 4, e109–16 (2015).

(29) Le, J. et al. Bayesian Estimation of Vancomycin Pharmacokinetics in Obese Children: Matched Case-Control Study. Clin Ther 37, 1340–51 (2015).

(30) Liu, T. et al. Population pharmacokinetics of vancomycin in Chinese pediatric patients Int J Clin Pharmacol Ther 55, 509–16 (2017).

(31) Moffett, B.S., Resendiz, K., Morris, J., Akcan-Arikan, A. & Checchia, P.A. Population Pharmacokinetics of Vancomycin in the Pediatric Cardiac Surgical Population. J Pediatr Pharmacol Ther 24, 107–16 (2019).

(32) Moffett, B.S., Ivaturi, V., Morris, J., Akcan Arikan, A. & Dutta, A. Population Pharmacokinetic Assessment of Vancomycin Dosing in the Large Pediatric Patient. Antimicrob Agents Chemother 63, (2019).

(33) Stockmann, C. et al. Population pharmacokinetics of intermittent vancomycin in children with cystic fibrosis. Pharmacotherapy 33, 1288–96 (2013).

(34) Zane, N.R. et al. A Population Pharmacokinetic Analysis to Study the Effect of Therapeutic Hypothermia on Vancomycin Disposition in Children Resuscitated From Cardiac Arrest. Pediatr Crit Care Med 18, e290–e7 (2017).

(35) Zhang, H. et al. Pharmacokinetic Characteristics and Clinical Outcomes of Vancomycin in Young Children With Various Degrees of Renal Function. J Clin Pharmacol 56, 740–8 (2016).

(36) Zhao, W. et al. Population pharmacokinetics and dosing optimization of vancomycin in children with malignant hematological disease. Antimicrob Agents Chemother 58, 3191–9 (2014).

(37) Villena, R., Gonzalez, C.A., Nalegach, M.E., Vasquez, A., Villareal, M. & Drago, M. [Therapeutic monitoring of intravenous vancomycin in a pediatric critical care unit]. Rev Chilena Infectol 31, 249–53 (2014).

(38) Acuna, C., Morales, J., Castillo, C. & Torres, J.P. [Pharmacokinetics of vancomycin in children hospitalized in a critical care unit]. Rev Chilena Infectol 30, 585–90 (2013).

(39) Gous, A.G., Dance, M.D., Lipman, J., Luyt, D.K., Mathivha, R. & Scribante, J. Changes in vancomycin pharmacokinetics in critically ill infants. Anaesth Intensive Care 23, 678–82 (1995).

(40) Giachetto, G.A., Telechea, H.M., Speranza, N., Oyarzun, M., Nanni, L. & Menchaca, A. Vancomycin pharmacokinetic-pharmacodynamic parameters to optimize dosage administration in critically ill children. Pediatr Crit Care Med 12, e250–4 (2011).

(41) Benner, K.W., Worthington, M.A., Kimberlin, D.W., Hill, K., Buckley, K. & Tofil, N.M. Correlation of vancomycin dosing to serum concentrations in pediatric patients: a retrospective database review. J Pediatr Pharmacol Ther 14, 86–93 (2009).

(42) Joo, S., Kim, M.S., Yang, J. & Park, J. Generative Model for Proposing Drug Candidates Satisfying Anticancer Properties Using a Conditional Variational Autoencoder. ACS Omega 5, 18642–50 (2020).

(43) Lim, J., Ryu, S., Kim, J.W. & Kim, W.Y. Molecular generative model based on conditional variational autoencoder for de novo molecular design. J Cheminform 10, 31 (2018).

(44) Matsuo, H. et al. Prognosis prediction of patients with malignant pleural mesothelioma using conditional variational autoencoder on 3D PET images and clinical data. Med Phys 50, 7548–57 (2023).

(45) Cuomo, S., Di Cola, V.S., Giampaolo, F., Rozza, G., Raissi, M. & Piccialli, F. Scientific Machine Learning Through Physics–Informed Neural Networks: Where we are and What’s Next. Journal of Scientific Computing 92, 88 (2022).

(46) Raissi, M., Perdikaris, P. & Karniadakis, G.E. Physics-informed neural networks: A deep learning framework for solving forward and inverse problems involving nonlinear partial differential equations. Journal of Computational Physics 378, 686–707 (2019).

(47) Diederik, P.K. & Max, W. Auto-Encoding Variational Bayes. eprint arXiv 1312.6114, (2013).

(48) Mingtian, Z., Peter, H. & David, B. Generalization Gap in Amortized Inference. eprint arXiv 2205.11640, (2022).

(49) van Groen, B.D. et al. Ontogeny of Hepatic Transporters and Drug-Metabolizing Enzymes in Humans and in Nonclinical Species. Pharmacol Rev 73, 597–678 (2021).

