## Supplemental materials for "Physiology-Informed Conditional Variational Autoencoder for Generating Pediatric Virtual Patients"

**Physiological data generations**

Physiological data corresponding to the observed NHANES data (age, sex, weight, height, and serum creatinine [SCR]) were generated using mechanistic models based on previously published reports (1, 2).

**Fat-free mass (FFM)**

Individual FFM (kg) was calculated based on the method described by O’Hanlon et al. (1) as follows:

$$FFM=FRFFM\times{FFM}_{adult}$$

FRFFM represents the fraction of fat-free mass (FFM) in pediatric subjects relative to the adult reference model (FFM_adult) reported by Janmahasatian et al (3). It is calculated by combining a baseline component (FFMIN), a neonatal/infant component (FFNEO), and a childhood component (FFKID), as follows:

$$FRFFM=FFMIN+FFNEO+FFKID$$

The derived individual FFM values corresponding to the observed NHANES data are shown below.

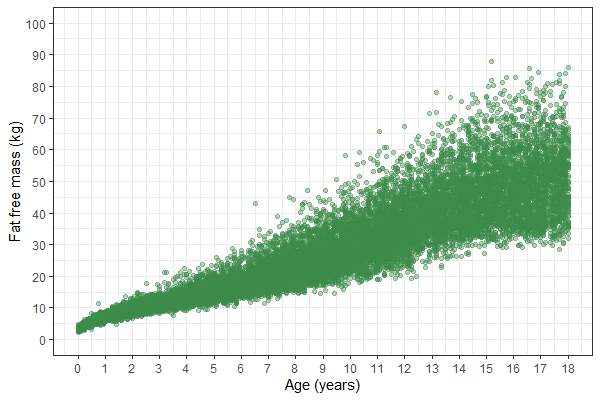

**Estimated glomerular filtration rate (eGFR) in subjects ≥ 12 years olds**

Individual eGFR (mL/min) was also calculated based on the method described by O’Hanlon et al.(1) as follows:

$$\mathrm{eGFR}=\frac{CPR}{SCR}$$

Where CPR is the creatinine production rate and SCR is the observed serum creatinine level from the NHANES database. NHANES used a modified Jaffe method for SCR assays through the 2015–2016 survey cycle. Beginning with the 2017–2018 cycle, an enzymatic method was used. Since no significant difference was observed between the two assay methods, the SCR values were used without adjustment. CPR was calculated as follows:

$$CPR={CPR}_{std}{\times F}_{size}\times F_{MAT,CPR}$$

Where CPR_std_ is the standard CPR in adults, F_size_ is the size factor, and F_MAT,CPR_ is the maturation factor for CPR.

**eGFR and SCR in subjects < 12 years olds**

For subjects under 12 years of age, SCR data were not available in the NHANES database. As a first step, individual eGFR was estimated as follows:

$$eGFR={GFR}_{std}{\times F}_{size}\times F_{MAT, PMA}\times F_{MAT,PNA}$$

Where GFR_std_ is the standard adult eGFR, F_size_ is the size factor, and F_MAT,PMA_ and F_MAT,PNA_ are maturation factors corresponding to postmenstrual age (PMA) and postnatal age (PNA), respectively. To reflect individual variability in renal function, inter-individual variability (IIV) was incorporated according to the estimates reported by O’Hanlon et al. Based on the calculated individual eGFR and CRP, SCR was derived as follows:

$$SCR=\frac{CPR}{\mathrm{eGFR}}$$

Individual eGFR and SCR values are shown below: purple dots for ages 0–12, and blue dots for ages 12–18.

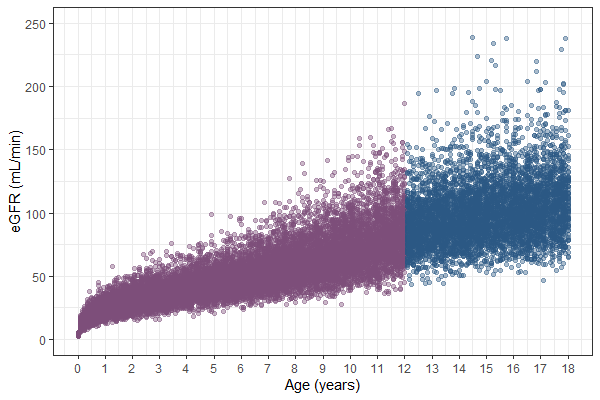

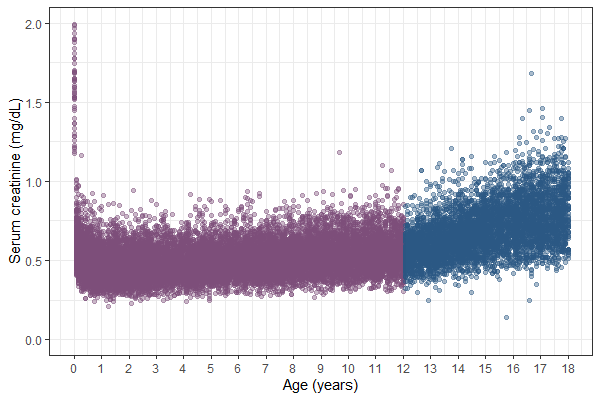

**Organ weights, Cardio output, and organ blood flows**

Mean organ weights (kg), cardiac output (L/min), and organ blood flows (mL/min/100 g organ weight) across age and sex were derived using mathematical equations reported by Chang et al (2). To account for differences in body size within the same age and sex groups, individual organ weights corresponding to subjects from the NHANES dataset were scaled by FFM for all organs except fat mass, and by total body weight for fat mass, as follows:

$$Organ weights =Mean organ weights \times\frac{Individual FFM or WT}{Standard FFM or WT}$$

Where the mean organ weights are predictions based on the mathematical equations by Chang et al., the individual FFM or total body weight (WT) values are taken from the NHANES dataset, and the standard FFM or WT values for each age and sex are derived using the equations by Chang et al. Also, individual cardio output was allometrically scaled by total body weight as follows:

$$Cardio Output =Mean Cardio Output\times\left( \frac{Individual WT}{Standard WT} \right)^{3/4}$$

Where the mean cardiac output is predicted using the mathematical equation by Chang et al., the individual WT values are taken from the NHANES dataset, and the standard WT values for each age and sex are derived using the equations by Chang et al (2).

Finally, individual organ blood flows (mL/min) were computed based on the corresponding organ weights. The resulting organ weights (kg), cardiac output (L/min), and organ blood flows (L/min) are presented below.

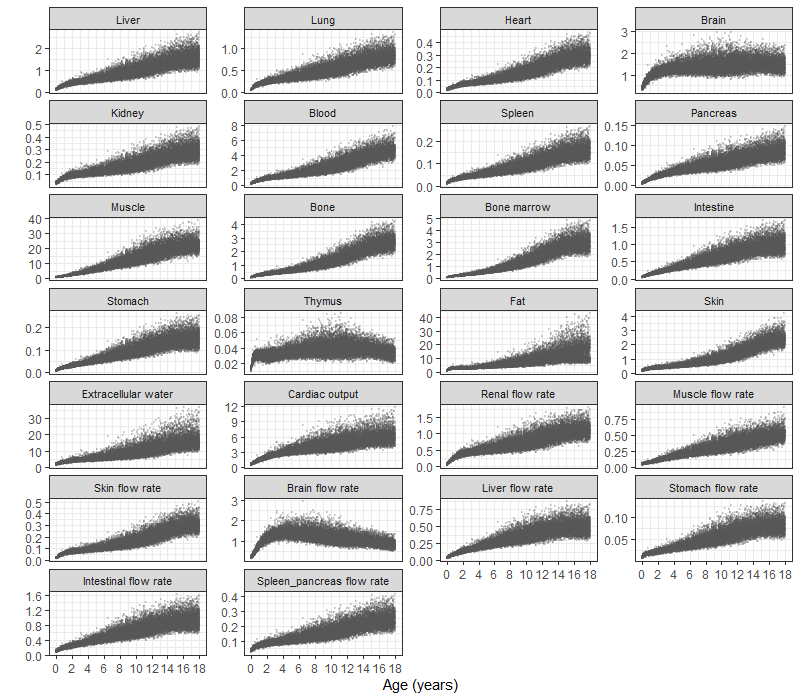

**Loss functions in the cVAE training**

The cVAE model (4) was trained using a composite loss function consisting of three components: a reconstruction loss, a Kullback–Leibler (KL) divergence term, and physiological constraints. Physiological constraints were implemented to penalize outputs in which the sum of organ weights exceeded total body weight, and the sum of organ blood flows exceeded cardiac output.

**Reconstruction loss**

The reconstruction loss (L_recon_) was defined as the mean squared error (MSE) between the input x and its reconstruction x̂, both computed in the normalized space:

$$L_{recon} =\mathbb{E} \left[ \left( x_{i}-\hat{x}_{i} \right)^{2} \right]$$

**KL divergence**

The KL divergence regularizes the approximate posterior q(z|x,c) toward a standard normal prior p(z) = N(0, I), promoting smoothness and disentanglement in the latent space.

$$L_{KL} =-\frac{1}{2}\mathbb{E}\left[ \sum_{j=1}^{D} \left( 1+\log\left( \sigma_{j}^{2} \right)-\mu_{j}^{2}-\sigma_{j}^{2} \right) \right]$$

Where μ_j_ and σ_j_ are the mean and standard deviation (obtained from the log-variance) of the approximate posterior q(z∣x,c) for the j-th latent dimension, and D is the dimensionality of the latent space, which was set to 2.

**Organ weight ratio loss**

To ensure physiological plausibility, this term penalizes outputs where the total organ weight exceeds total body weight:

$$L_{organ\_ratio} =\mathbb{E}\left[ max\left( 0,\frac{{\sum_{k} \hat{x}}_{k}^{pyhs}}{\hat{x}_{WT}^{pyhs}+\varepsilon}-1 \right) \right]$$

Where $\hat{x}_{k}^{pyhs}$denotes the reconstructed weight of organ k, and $\hat{x}_{WT}^{pyhs}$denotes the reconstructed body weight, both in physical units (kg). The total organ set includes: Liver, Lung, Heart, Brain, Kidney, Blood, Spleen, Pancreas, Muscle, Bone, Bone marrow, Intestine, Stomach, Thymus, Fat, and Skin. ε is a small constant to avoid division by zero.

**Cardiac output ratio loss**

This term penalizes outputs where the sum of organ-specific blood flows exceeds the total cardiac output:

$$L_{co\_ratio} =\mathbb{E}\left[ max\left( 0,\frac{{\sum_{k} \hat{x}}_{blood flow\_k}^{pyhs}}{\hat{x}_{co}^{pyhs}+\varepsilon}-1 \right) \right]$$

Where $\hat{x}_{blood flow\_k}^{pyhs}$denotes the reconstructed blood flow to organ k, and $\hat{x}_{co}^{pyhs}$denotes the reconstructed cardiac output, both in physical units (L/min). The total blood flow set includes: Liver, Brain, Kidney, Muscle, and Skin. Blood flows to gastrointestinal organs were excluded to prevent double-counting, as these flows are physiologically routed via the portal vein and incorporated into liver blood flow. ε is a small constant to avoid division by zero.

**Total loss**

The total loss is defined as a weighted sum of the above components:

$$L_{total} =\lambda_{recon}\cdot L_{recon}+\lambda_{KL}\cdot L_{KL}+\lambda_{organ\_ratio}\cdot L_{organ\_ratio}+\lambda_{co\_ratio}\cdot L_{co\_ratio}$$

where each λ represents a tunable weight for the corresponding loss term. In the final cVAE model, $\lambda_{KL}$ was 0.0008, while all other loss weights were set to 1.0.

**Implementation of the Pediatric PBPK Model**

The pediatric PBPK model was implemented in R using the mrgsolve package (version 1.5.2), based on the framework described by Poulin et al (5). The model included compartments for adipose tissue, bone, brain, gut, heart, kidney, liver, lung, muscle, skin, spleen, and the rest of the body. Organ volumes and organ-specific blood flows were used as inputs from the cVAE outputs. The rest-of-body compartment was defined as the remaining fraction of total body weight after accounting for all explicitly modeled organs. Because blood flow estimates for adipose tissue, heart, and the rest-of-body compartment were not available, the remaining cardiac output was distributed to these compartments according to adult physiological flow ratios.

**Development of the Vancomycin PBPK Model in Pediatric Patients**

Tissue-to-plasma partition coefficients (Kp) for vancomycin were predicted using equations for drugs that primarily distribute into the extracellular space (6), assuming a plasma unbound fraction of 0.45. Renal clearance and a global Kp scaling factor were estimated by least-squares optimization using the derivative-free NEWUOA algorithm (minqa R package, version 1.2.8), based on reported mean concentration–time profiles from pediatric patients aged 2.6 days to 7.6 years (7), with age- and body weight–matched cVAE-generated organ volumes and blood flows used as model inputs.

**PBPK simulations using cVAE generated physiology data**

Latent vectors (z1 and z2) were sampled from a standard normal distribution (mean 0, SD 1) and assigned age (0–18 years) and sex. The decoded physiological parameters were used as inputs to the vancomycin PBPK model. The area under the concentration–time curve over 24 hours (AUC₀–₂₄) and the maximum concentration (Cmax) following a single 10 mg/kg dose administered as a 1-hour infusion were simulated and compared with reported pediatric observations across the full pediatric age range, including special populations such as patients with renal impairment and obesity.

Literature AUC values across ages and populations were derived from reported clearance estimates in population pharmacokinetic studies by calculating AUC₀–₂₄ for a 10 mg/kg dose using typical age-, body weight–, and covariate-adjusted clearance values reported in each study. Reported Cmax values were proportionally normalized to a 10 mg/kg dose. These reference data were compiled from review articles (8, 9) and original studies, as summarized in **Tables S2 and S3**.

**References**

(1)	O'Hanlon, C.J., Holford, N., Sumpter, A. & Al-Sallami, H.S. Consistent methods for fat-free mass, creatinine clearance, and glomerular filtration rate to describe renal function from neonates to adults. *CPT Pharmacometrics Syst Pharmacol* **12**, 401-12 (2023).

(2)	Chang, H.P., Kim, S.J., Wu, D., Shah, K. & Shah, D.K. Age-Related Changes in Pediatric Physiology: Quantitative Analysis of Organ Weights and Blood Flows : Age-Related Changes in Pediatric Physiology. *AAPS J* **23**, 50 (2021).

(3)	Janmahasatian, S., Duffull, S.B., Ash, S., Ward, L.C., Byrne, N.M. & Green, B. Quantification of lean bodyweight. *Clin Pharmacokinet* **44**, 1051-65 (2005).

(4)	Sohn, K., Lee, H. & Yan, X. Learning Structured Output Representation using Deep Conditional Generative Models. In eds. Cortes, C., Lawrence, N., Lee, D., Sugiyama, M. and Garnett, R.).

(5)	Poulin, P. & Theil, F.P. Prediction of pharmacokinetics prior to in vivo studies. II. Generic physiologically based pharmacokinetic models of drug disposition. *J Pharm Sci* **91**, 1358-70 (2002).

(6)	Poulin, P. & Theil, F.P. Prediction of pharmacokinetics prior to in vivo studies. 1. Mechanism-based prediction of volume of distribution. *J Pharm Sci* **91**, 129-56 (2002).

(7)	Schaad, U.B., McCracken, G.H., Jr. & Nelson, J.D. Clinical pharmacology and efficacy of vancomycin in pediatric patients. *J Pediatr* **96**, 119-26 (1980).

(8)	Aljutayli, A., El-Haffaf, I., Marsot, A. & Nekka, F. An Update on Population Pharmacokinetic Analyses of Vancomycin, Part II: In Pediatric Patients. *Clin Pharmacokinet* **61**, 47-70 (2022).

(9)	Akunne, O.O., Mugabo, P. & Argent, A.C. Pharmacokinetics of Vancomycin in Critically Ill Children: A Systematic Review. *Eur J Drug Metab Pharmacokinet* **47**, 31-48 (2022).
